# Age-sex specific prevalence and incidence of antidepressant prescribing in the Nordic countries: a systematic review

**DOI:** 10.64898/2025.12.04.25341462

**Authors:** Marcel Ballin, Paulina Tuvendal, Carola Bardage, Mats Persson, Mats Talbäck, Rickard Ljung

## Abstract

Marked differences in antidepressant prescribing have been reported across the Nordic countries, but existing reviews are based on older data and focus mainly on young people. We conducted an updated systematic review of antidepressant prescribing across all age groups in the Nordic countries. We searched Embase, PubMed, and Web of Science for observational studies reporting prevalence and/or incidence of antidepressant prescribing in general populations in the Nordic countries, published from 1 January 2000 to 1 October 2025. From 2728 screened articles, 30 studies with prescribing data from 1994–2021 were included. The rate of antidepressant prescribing was higher in Sweden compared to Denmark and Norway, and these differences increased in recent years, mainly due to large increases in Sweden. Findings were consistent across all age groups and both sexes, but were especially pronounced among children and adolescents, and particularly girls. The most recent data showed a 2–5 times higher prevalence and incidence of antidepressant prescribing among Swedish girls and boys as compared to their Danish and Norwegian peers, including relevant differences in absolute measures. The prevalence pattern was similar in adults, and while relative differences were smaller there were large absolute differences. Comparisons with Finland and Iceland were not feasible due to outdated data. The rate of antidepressant prescribing is consistently higher in Sweden compared to in Denmark and Norway, and these differences between countries have increased markedly over time. The pattern is observed across all age groups and both sexes but is especially strong among young females.

## Introduction

Psychiatric disorders, such as depressive, anxiety, and obsessive-compulsive disorders, account for a large proportion of global disease burden and are associated with reduced life expectancy and significant economic burden [1–4]. The Nordic countries, which consist of Denmark, Finland, Iceland, Norway, and Sweden, have a high psychiatric morbidity and mortality burden [1, 4, 5], as well as a high consumption of antidepressive medications used in the treatment of several of these disorders [6]. However, despite the many similarities between the Nordic countries, such as comparable populations, healthcare- and drug reimbursement systems [7], as well as burden of psychiatric disorders such as depression [5], emerging evidence indicates increased variations across the countries in the prescribing of antidepressants [8].

A recent systematic review of antidepressant prescribing in the Nordic countries found a much higher prescribing in Sweden compared to in Denmark and Norway [8]. However, the review focused on children and adolescents and was based on older data, as the most recent prevalence data was from 2017 and the most recent incidence data from 2013. Whether the gap in prescribing may have narrowed or even widened in recent years remains unclear, but it may be hypothesized that prescribing could have continued to increase in recent years [9], alongside a worsening in mental health [10]. An updated synthesis of contemporary, population-wide evidence, especially with age- and sex-specific estimates, could yield valuable insights on recent trends. Therefore, we conducted a systematic review of the prevalence and incidence of antidepressant prescribing across all age groups in the Nordic countries, with a focus on time trends in age-sex specific estimates.

## Methods

This systematic review was conducted within the scope of a government investigation targeting antidepressant prescribing in Sweden compared to the other Nordic countries, which was assigned to the Swedish Medical Products Agency on 16 December 2024 on behalf of the Government Offices of Sweden, Ministry of Social Affairs. The protocol for this review was pre-registered (Prospero CRD420250651803) [11], and the report follows the PRISMA guidelines [12].

### Eligibility criteria

Eligible studies were observational studies written in English or Scandinavian languages, published since 2000 and reporting yearly prevalences and/or incidences of antidepressant prescribing in the general population in the Nordic countries, with antidepressant prescribing defined using Anatomical Therapeutic Chemical classification system code N06A. We excluded studies meeting any of the following criteria: (i) not focusing on antidepressant prevalence/incidence per se, (ii) conducted in non-population-based samples (e.g. clinical samples), (iii) only reporting cumulative estimates, (iv) relying on self-reported drug use.

### Literature search strategy

An information specialist at the Swedish Medical Products Agency developed the literature search strategy (supplemental file 1) in collaboration with three reviewers (MB, PT, and RL). Literature searches were carried out in Embase, PubMed, and Web of Science, searching for articles published until 4 April 2025, with an updated search for articles published until 1 October 2025, using keywords like “antidepressants”, “prevalence”, and “incidence”. Both free text terms and controlled terms were used. Boolean operators were applied to combine terms and proximity operators were used when appropriate. The literature search was supplemented by backward and forward citation searching of included articles.

### Study selection and data extraction

The articles were imported into Rayyan systematic review software [13]. After duplicates removal, one reviewer (MB) screened all articles at the title level and two reviewers (MB and PT) independently screened all remaining articles at the abstract and full-text level. Disagreements were discussed and conflicts were resolved by consulting a third reviewer (RL), until consensus was reached. Data extraction of all included studies was performed by one reviewer (MB) and double-checked by a second reviewer (MT). The data were extracted into Microsoft Excel and included bibliometric information, study characteristics (ie, study period, country, population size, age, proportion of female participants, antidepressant definition), and outcome data (ie, yearly prevalence and/or incidence of prescribed antidepressants). We contacted the authors of articles in which relevant data were not sufficiently reported, such as age-sex specific estimates. Estimates were harmonized to be expressed as proportions per 1000 population/person-years.

### Study quality

Study quality of all included studies was assessed independently by two reviewers (MB and PT) using the Joanna Briggs Institute Critical Appraisal Checklist for Studies Reporting Prevalence Data [14]. This tool assigns a score ranging from 0 (lowest quality) to 9 (highest quality) based on the following domains: appropriateness of the sample frame; recruitment procedure; adequacy of the sample size; description of participants and setting; coverage of the identified sample; validity of the method used to identify antidepressants; reliability of the method used to identify antidepressant; adequacy of statistical analyses; and response rate. One point per domain is assigned.

## Results

### Study selection

The literature search generated a total of 4697 records. After duplicates removal, 2728 unique articles remained and were screened at the title and abstract level against eligibility criteria. Of these, 54 were subject to screening at full-text level. From these articles, 24 were excluded (supplemental table 1) while 30 met the inclusion criteria. Backward and forward citation searching did not yield additional articles. Thus, a total of 30 articles were included in the review (figure 1).

**Figure 1.**
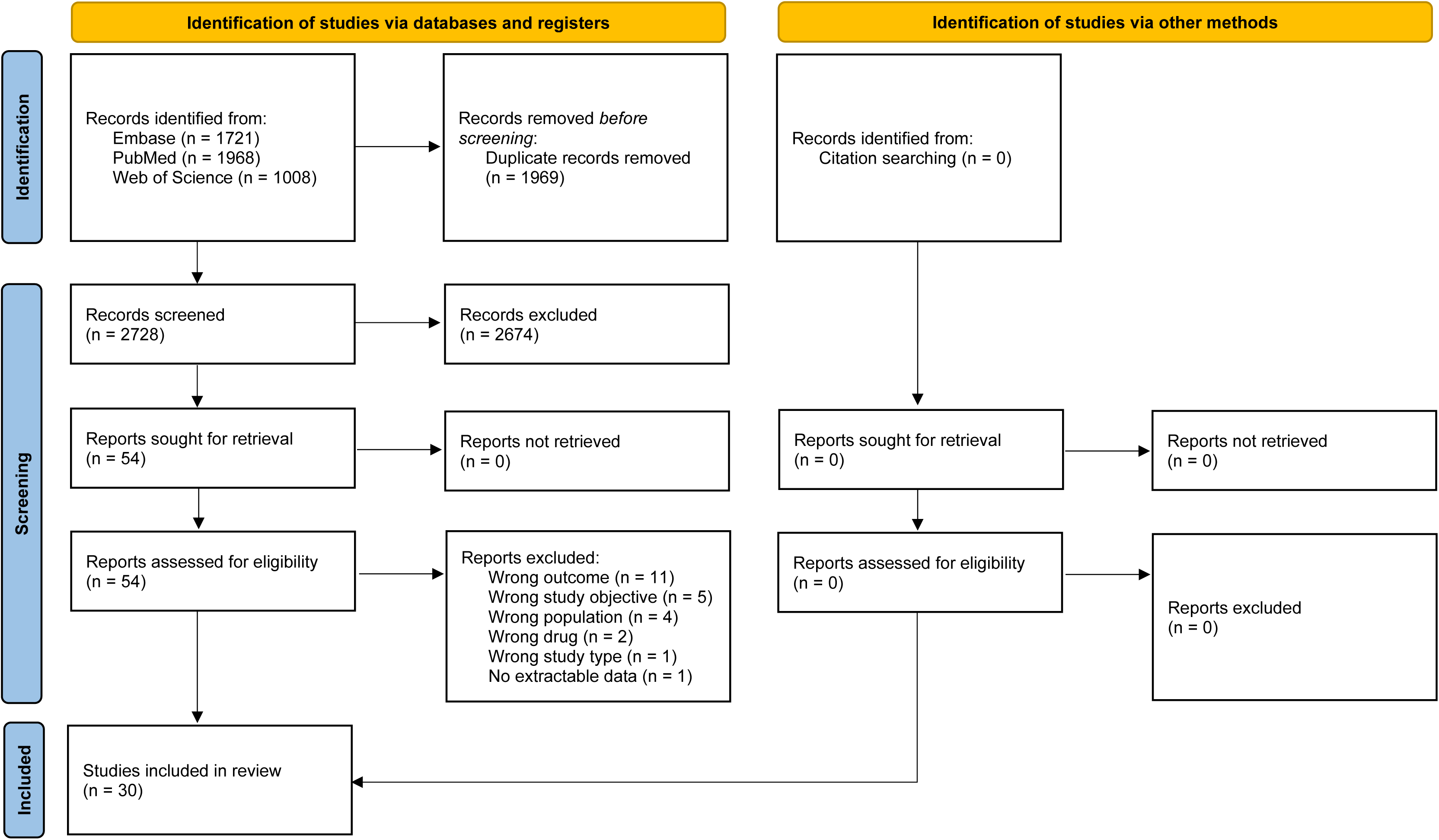
**PRISMA flowchart of the study selection.**

### Study characteristics and quality

The 30 studies (table 1) reported estimates from 1994 to 2021, and covered Denmark (n=13, of which 1 also included Greenland), Finland (n=5), Iceland (n=1), Norway (n=10), and Sweden (n=9) [15–44]. Prevalence of prescribed antidepressants was reported in 26 studies and incidence in 11 studies. Ages in the studies varied and sometimes comprised both children and adults, but almost half of the studies (n=14) exclusively targeted people aged <20 years, while almost a quarter (n=7) focused on adults. Twenty-nine studies used large-scale nationwide or regional prescription/medical databases, whereas one included review of medical records. Twenty-eight considered all antidepressants, whereas two studies were restricted to selective serotonin reuptake inhibitors.

In the study quality assessment, 29 studies received a score of ≥7, and one study received a score of 5 (supplemental table 2). The individual domains for which almost all studies (n=28) did not score a point were the statistical analysis (due to lack of complete reporting of outcome data), and the description of study participants and setting (n=27).

### Prevalence of antidepressant prescribing

The 26 studies on prevalence covered Denmark (n=11, of which 1 also covered Greenland), Finland (n=4), Iceland (n=1), Norway (n=8), and Sweden (n=8) (supplemental table 3) [15–39, 44]. From these, 12 studies reported age-sex specific prevalences across several years [16–19, 21, 23, 27, 28, 33, 34, 37, 44].

#### Children and adolescents

Overall, the prevalence of antidepressant prescription was higher among girls than boys and increased with age. Based on the most recent available data, comparisons across countries showed a roughly three times higher prevalence among Swedish boys and girls aged 15–19 years compared to their Danish and Norwegian peers, and about three to five times higher among those aged 10–14. Key findings in terms of temporal trends are described below and are illustrated graphically in figure 2 and figure 3.

**Figure 2.**
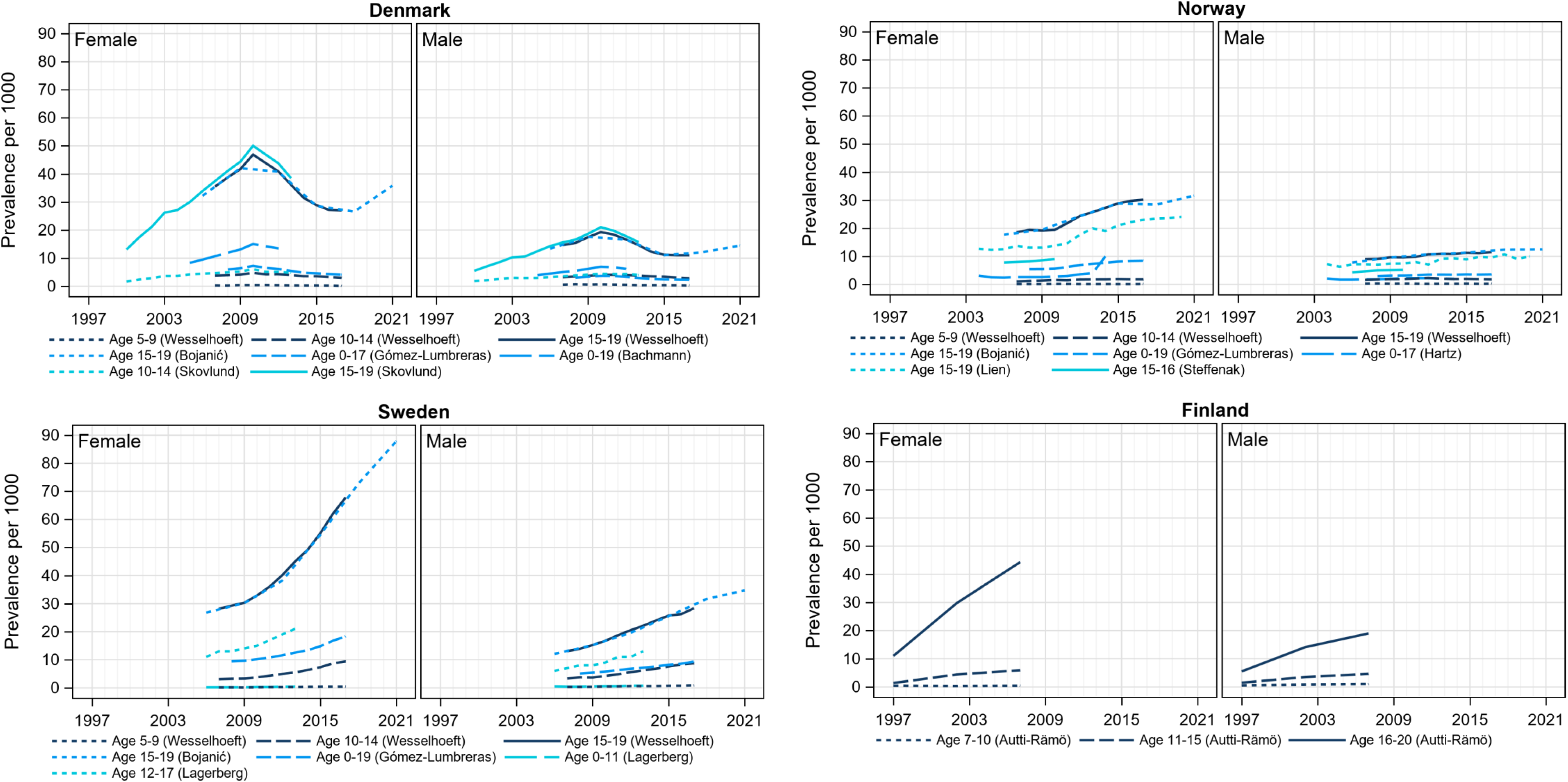
Trends in age-sex specific prevalence of prescribed antidepressants among children and adolescents in Denmark, Finland, Norway, and Sweden. Presented estimates were selected based on comparable age-sex groups. Remaining estimates were omitted for clarify as they are described in supplemental table 3.

**Figure 3.**
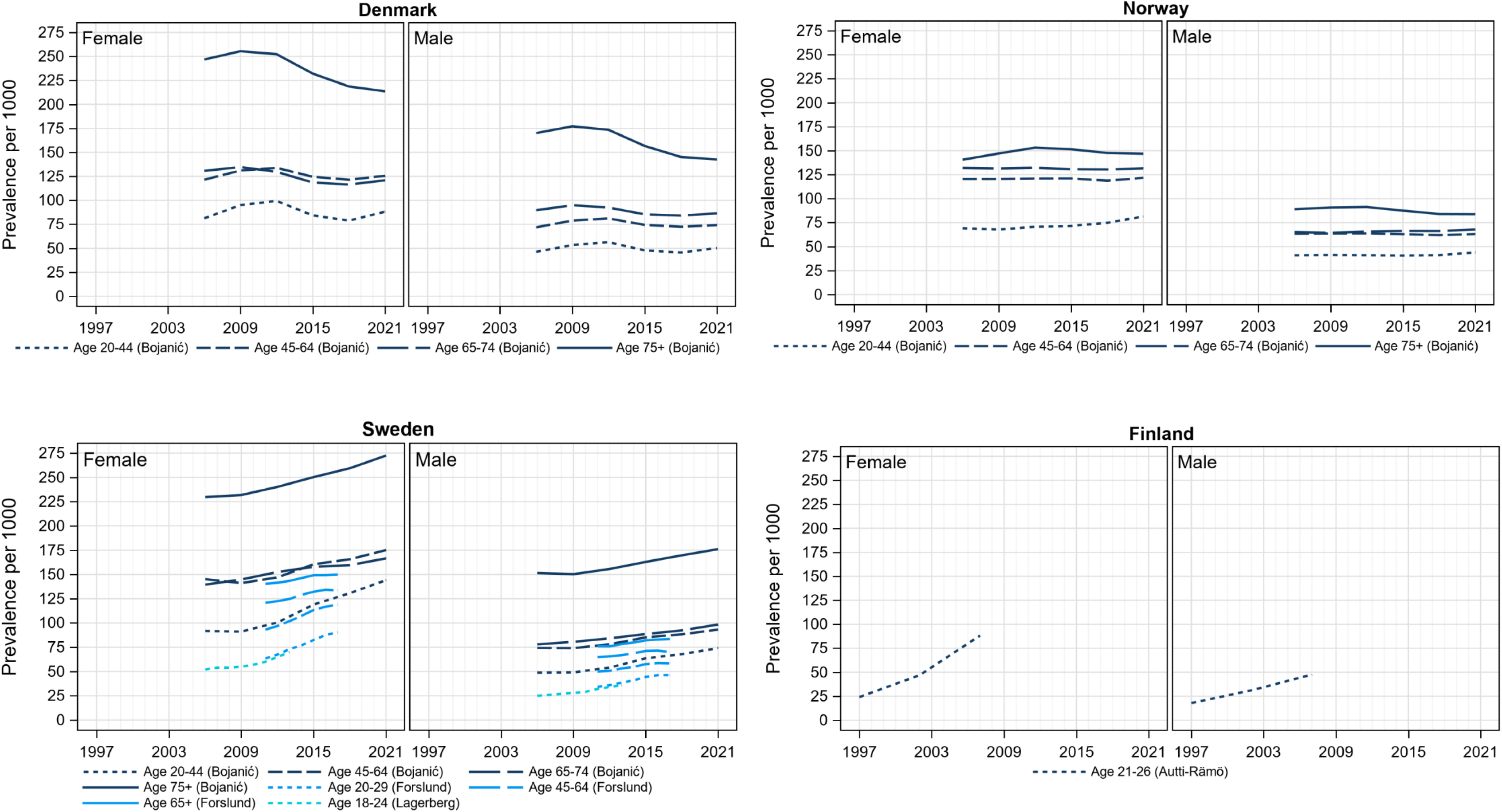
**Trends in age-sex specific prevalence of prescribed antidepressants among adults in Denmark, Finland, Norway, and Sweden.**

**Figure 4.**
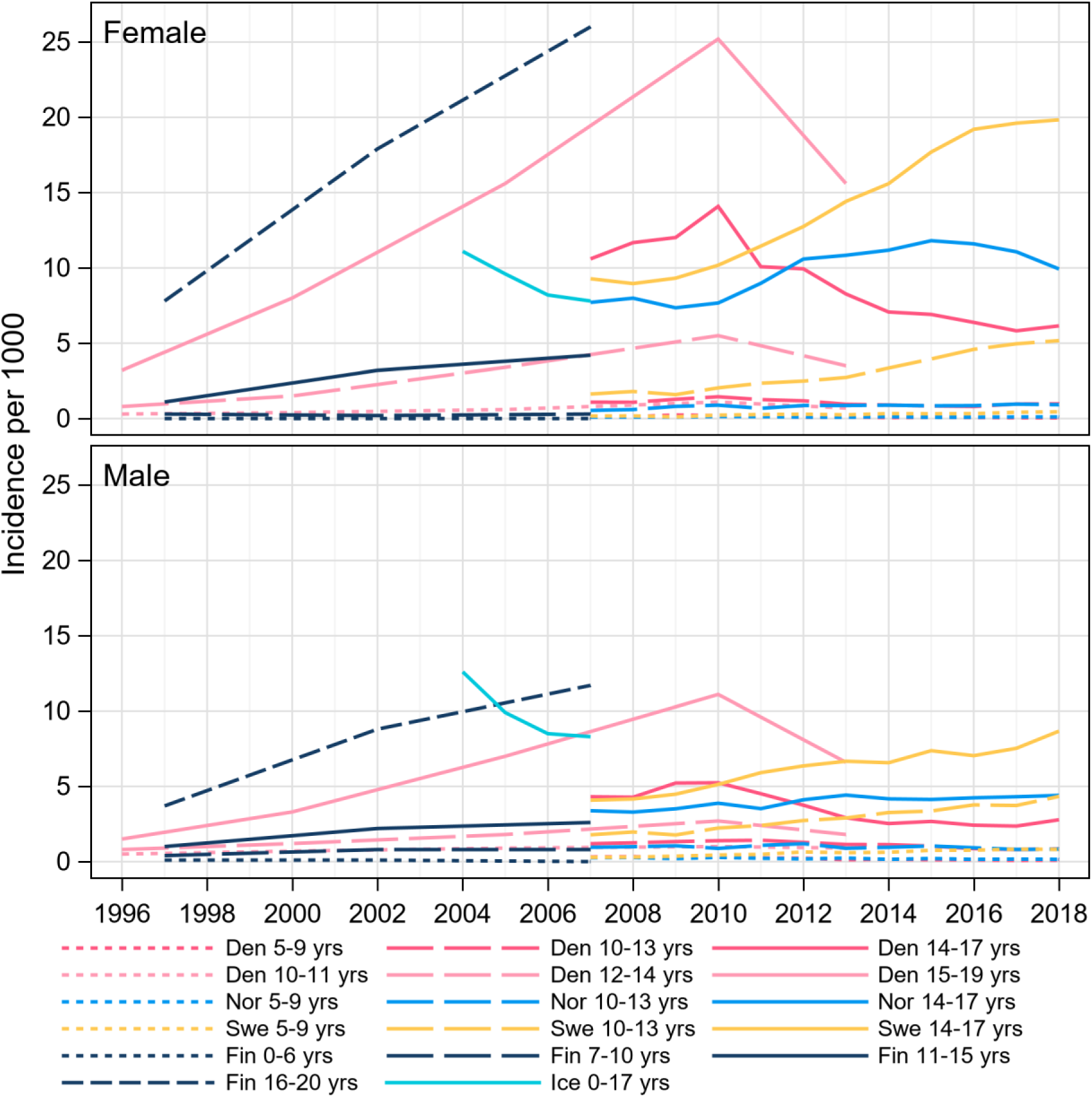
Trends in age-sex specific incidence of prescribed antidepressants among children and adolescents in Denmark, Finland, Iceland, Norway, and Sweden. Presented estimates were selected from four studies,^16,33,39,40^ including estimates from comparable age-sex groups. Remaining estimates were omitted for clarity as they are described in supplemental table 4.

In Denmark, estimates between 2000 and 2021 were reported [17, 18, 21, 33, 37]. The prevalence increased until a peak around 2010, after which it began declining. Among girls aged 10–14, one study reported a prevalence of 1.7/1000 in 2000, which increased to 6.1/1000 in 2010, and decreased to 4.8/1000 in 2013.[33] In another study, the prevalence decreased further to 3.0/1000 in 2017 [37]. Among boys aged 10–14, the prevalence increased from 1.9/1000 in 2000 to 4.4/1000 in 2010, after which it decreased to 4.0/1000 in 2013,[33] and further to 2.8/1000 in 2017 [37]. Among girls aged 15–19, the prevalence increased from 13.1/1000 in 2000 to 50.0/1000 in 2010, and decreased to 38.5/1000 in 2013 [33], and further to 27.0/1000 in 2017 [37]. Among boys aged 15–19, the prevalence increased from 5.5/1000 in 2000 to 21.0/1000 in 2010, after which it declined to 15.7/1000 in 2013 [33], and further to 11.1/1000 in 2017 [37]. In another study, the prevalence increased slightly in the most recent years among those aged 15–19 [18]. Among boys, the prevalence increased from 11.1/1000 to 14.5/1000 between 2015 and 2021 and among girls from 26.6/1000 to 35.8/1000 between 2018 and 2021.

In Sweden, estimates between 2006 and 2021 were reported [18, 21, 27, 37]. The prevalence increased over time with no evidence of leveling off. Among girls aged 10–14, the prevalence increased from 3.0/1000 to 9.4/1000 between 2007 and 2017, and among boys from 3.4/1000 to 8.7/1000 [37]. Among girls aged 15–19, the prevalence between 2006 and 2021 increased from 26.8/1000 to 87.9/1000 and among boys from 12.1/1000 to 34.7/1000 [18].

In Norway, estimates between 2004 and 2021 were reported [18, 21, 24, 28, 34, 37]. The prevalence among girls aged 15–19 increased from 17.7/1000 to 31.6/1000 between 2006 and 2021, and among boys from 7.8/1000 to 12.5/1000. Among those aged 10–14 years the prevalence remained more stable.

In Finland, one study reported prevalence between 1997 and 2007 [16]. Among those aged 11–15, the prevalence in boys increased from 1.5/1000 to 4.6/1000, and in girls from 1.4/1000 to 5.9/1000. Among those aged 16–20, the prevalence in boys increased from 5.5/1000 to 19.0/1000, and in girls from 11.0/1000 to 44.3/1000.

#### Adults

Four of the studies reported age-sex specific prevalence among adults over time [16, 18, 19, 27]. Based on the most recent available data, comparisons across countries showed a roughly one and a half times higher prevalence among Swedish men and women aged 20–44 compared to their Danish and Norwegian peers, and about 1.3-2 times higher prevalence among those aged ≥75 years. Key findings in terms of temporal trends are described below.

In Norway, the prevalence remained relatively stable between 2006 and 2021. Among men aged 20–44 years, the prevalence was 40.9/1000 in 2006 and 44.0/1000 in 2021, and among women of the same age it was 69.2/1000 in 2006 and 81.5/1000 in 2021 [18]. Corresponding estimates for those aged ≥75 years were 89.0/1000 to 83.8/1000 among men, and 140.6/1000 to 146.8/1000 among women [18].

In Denmark, the prevalence estimates were comparable to those in Norway, and changes over time were mostly stable. The exception was older adults aged ≥75, for which the prevalence was higher compared to their Norwegian peers, although declining between 2006 and 2021 among both men (from 170.1/1000 to 142.6/1000) and women (from 246.8/1000 to 213.6/1000) [18].

In contrast, the prevalence in Sweden was higher among both men and women across all age groups compared to in Denmark and Norway, and it increased over time. For example, among women aged 20–44, the prevalence increased from 91.8/1000 in 2006 to 144.1/1000 in 2021, and among men from 48.4/1000 to 74.1/1000 [18]. Among those aged ≥75 years, the prevalence among women increased from 229.7/1000 to 272.4/1000, and among men from 151.5/1000 to 176.1/1000 [18].

### Incidence of antidepressant prescribing

From the 11 studies that reported incidence [16, 20, 24, 29, 30, 33, 39–43], we were unable to extract relevant data from one study [24], leaving 10 studies covering Denmark (n=4), Finland (n=3), Iceland (n=1), Norway (n=2), and Sweden (n=2) (supplemental table 4) [16, 20, 29, 30, 39–43]. Five of these reported age-sex specific incidences over several years [16, 33, 39, 40, 43].

#### Children and adolescents

Based on the most recent available data, comparisons across countries showed a roughly four times higher incidence among Swedish boys and girls aged 10–13 compared to their Danish and Norwegian peers, and about two to three times higher incidence among those aged 14–17. Key findings in terms of temporal trends are described below and illustrated graphically in figure 3.

One study from Denmark reported on incidence between 1996 and 2013 [33]. In 1996, the incidence was lower in the younger age groups compared to older, but whereas the incidence declined over time in the older age groups, increases were seen in the younger age groups, particularly among females. However, the incidences declined after 2010, similar as for prevalence. For example, among girls aged 15–19, the incidence increased from 3.2/1000 to 15.6/1000 between 1996 and 2013 but decreased from the peak of 25.2/1000 in 2010. Another study reported incidences between 2007 and 2018 in Denmark, Norway, and Sweden [40]. Again, in Denmark, the incidence increased during the first few years after 2007 but declined after around 2010 [40]. Among girls aged 10–13, the incidence dropped from 1.45/1000 to 0.98/1000 between 2010 and 2018, and among boys it dropped from 1.42/1000 to 0.85/1000 between 2011 and 2018 [40]. Among girls aged 14–17, the incidence dropped from 14.09/1000 to 6.15/1000 between 2010 and 2018, and from 5.24/1000 to 2.78/1000 among boys [40].

In contrast, incidences in Sweden increased markedly throughout the study period. For example, among boys aged 14–17, the incidence increased from 4.08/1000 to 8.67/1000 between 2007 and 2018 [40]. Among girls of the same age, it increased from 9.28/1000 to 19.83/1000 [40]. Even larger increases were observed among those aged 10–13, where the incidence increased from 1.78/1000 to 4.33/1000 among boys, and from 1.63/1000 to 5.17/1000 among girls [40].

In Norway, incidences were generally more stable, but among girls aged 14–17 it increased from 7.71/1000 to 9.93/1000 between 2007 and 2018, although declining from a peak of 11.81/1000 in 2015 [40].

The studies from Finland and Iceland dated further back in time and covered a shorter time frame. In Finland [16], the incidence between 1997 and 2007 increased from 1.0/1000 to 2.6/1000 among boys 11–15 years and from 3.7/1000 to 11.7/1000 among boys 16–20 years. In girls, the incidence increased from 1.1/1000 to 4.2/1000 among those 7–10 years and from 7.8/1000 to 26.0/1000 among those 16–20 years. In Iceland [39], the incidence among boys 0–17 years fell from 12.6/1000 in 2004 to 8.3/1000 in 2007, and from 11.1/1000 to 7.8/1000 among girls.

#### Adults

There were limited data on changes in age-sex specific incidence in adults. In Denmark, the incidence among men and women aged 20–34 increased between 1996 and 2013, with a peak in 2010, whereas among men and women aged 35–49, the incidence in 2013 was roughly similar to or even lower than in 1996 [33]. Another study in Denmark examined the incidence among older adults, reporting a stable incidence between 2016 and 2018 among individuals aged ≥65 years, in both men (27.4/1000 to 25.7/1000) and women (36.8/1000 to 33.4/1000) [43]. In Finland, the incidence increased from 1997 to 2007 in both men (10.8/1000 to 23.8/1000) and women (14.2/1000 to 36.9/1000) aged 21–26 years [16].

There were no data on trends in the incidence among adults in either Norway or Sweden, with only two studies reporting incidence during a single year [29, 42]. In Norway, the incidence in 2008 among adults 20–39 years was 8.5/1000 in men and 12.7/1000 in women, which is about three times lower than their Finnish peers (ie, aged 21–26) in 2007 described above [42]. In Sweden, the incidence in 2010 in the total male population was 15.4/1000 and 24.7/1000 in the female population [29].

## Discussion

This systematic review confirmed large differences in antidepressant prescribing between the Nordic countries, with a higher prescribing in Sweden compared to Denmark and Norway. These differences between countries have amplified during recent years and are seen across all age groups and both sexes, but more pronounced among children and adolescents, and particularly girls.

A key finding of this review is that it revealed a growing divergence in prescribing during recent years, where in Denmark and Norway there has been either a stable, less pronounced, or even declining prescribing, as opposed to a continuous increase in Sweden. A recent review came to similar conclusions but only covered prevalence data until 2017 and incidence data until 2013 [8]. Our review confirms that the gap has further widened in recent years. As such, based on the most recent data, the prevalence of antidepressant prescribing among Swedish 15–19-year-olds was about three times higher, and among 10–14-year-olds up to five times higher, than their Danish and Norwegian peers. The incidence was also about four times higher among Swedish boys and girls aged 10–13 compared to their Danish and Norwegian peers, and about two to three times higher among those aged 14–17. While the differences in absolute terms were relatively small among those aged 10–14, the differences were larger among those aged 15–19. For example, the difference in prevalence when comparing Sweden to Denmark and Norway was approximately 52 to 56/1000 among girls and approximately 20 to 22/1000 among boys. Additionally, whereas the previous review focused exclusively on young people, our review suggests that differences between countries are evident also in adults. Although relative differences were smaller than for children and adolescents, there were large absolute differences. For example, among women aged 20–44 years the difference in prevalence was approximately 56 to 62 per 1000, and among women aged 75 years and older the difference was approximately 125 to 129 per 1000.

The reason for the shifts and variations in prescription patterns is likely multifactorial and influenced by several measurable, partly measurable, and unmeasurable factors. We cannot determine any causes for this since our review was descriptive in its nature, but factors such as drug reimbursement, licensing, and treatment recommendations are unlikely to play a major role in explaining the differences between countries. Denmark, Norway and Sweden have similar drug reimbursement systems [7], licensing status of antidepressive medications, and treatment guidelines. Yet, it should be noted that it is challenging to compare treatment guidelines since definitions (disease severity versus presence/absence of diagnosis) and guideline developers (governmental authorities, professional associations, etc.) may vary. In any case, our findings shed light on the importance of further analytical studies that seek to address potential causes for the shifts and variations in prescribing patterns, some of which are exemplified below.

One aspect could be to examine whether there are variations between countries in compliance with treatment guidelines. Even if the content of the guidelines is overall similar, it cannot be ruled out that the compliance differs, potentially because of variations in and/or clinician treatment preference, and availability and out-of-pocket costs of pharmacological versus non-pharmacological treatment. In Sweden for example, the National Board of Health and Welfare has stated that the availability of psychotherapy in the treatment of depression and anxiety disorders needs to increase [45]. Further, since the prevalence measure can be challenging to interpret as it is dependent on both the incidence and treatment duration, studies could seek to address whether there have been changes over time in treatment duration in the Nordic countries [40], which then could be taken into account when interpreting the differences in prevalence across countries.

It may also be valuable to examine whether there are differences between countries in terms of underlying psychiatric disorders where antidepressant medication is indicated, and if this could partly explain the difference in prescribing [46, 47]. Other factors that could be considered are more structural ones, such as the role of different care pathways and staff availability. For example, Sweden has been reported to have a higher availability of child and adolescent psychiatric specialists [48], and Swedish residents can use self-referral to specialist care, where the majority of antidepressants to children and adolescents are prescribed [40]. Thus, it is possible that part of the higher prescribing among young people in Sweden may relate to better access to qualified, specialist care. Finally, although difficult to study, cultural factors may be involved. For example, a study from Denmark suggested that the downward trend in incidence from around 2010 was the result of population skepticism towards antidepressant use as a result of increased media attention at this time, mainly revolving around side effects [49]. Collectively, the findings of this review show that analytical studies are warranted to understand the causes for the observed prescription patterns both within and between the Nordic countries. This would be important for evaluating equality of care and to ensure alignment with evidence-based practice. These insights may ultimately inform safer prescribing, more efficient resource allocation, and improved mental health care.

### Limitations and strengths

This work has some limitations that should be considered. First, we did not search grey literature for potential articles which means that we cannot rule out a risk of publication bias in this review. At the same time, this approach likely reduced the risk of including low-quality studies. Second, there were not studies from all the Nordic countries reporting estimates from the same year and with comparable age-sex stratification to produce a pooled estimate. In any case, this does not affect the implications of this review since the focus was to describe variations between countries and not the overall prescribing. Third, our review showed that there was much less data on antidepressant prescribing in adults compared to in children and adolescents, particularly regarding incidence, and this should be the target of future studies. Fourth, recent data on antidepressant prescribing in Finland and Iceland were lacking, and we only identified a very small number of studies with largely historical data. This prohibited us from performing comparisons with these countries, and we urge researchers to conduct and publish up-to-date analyses of antidepressant prescribing in these countries. Ideally, a large, multinational study including data from all Nordic countries and with harmonized definitions would be highly valuable. Fifth, because our review was restricted to studies conducted in the general population, and without data on the indication for treatment, the findings may not be generalized to certain segments of the population. Thus, it is possible that the temporal trends may differ between subgroups, such as depending on socioeconomic factors and comorbidities, and this should be further investigated.

Strengths of this review include systematic searches by an information specialist in three databases, the inclusion of about threefold the number of studies compared to the most recent similar review, the inclusion of studies on both prevalence and incidence across all ages, and the detailed breakdown of the data compared to the previous review, allowing for clearer insights into subgroup differences.

## Conclusions

In summary, this systematic review highlights a growing divergence in antidepressant prescribing between the Nordic countries, particularly in children and adolescents, and especially girls. These findings carry both clinical implications and guidance for future research, underscoring the need for analytical studies targeting potential reasons for the higher and increasing prescribing in Sweden, as well as updated prescribing studies in Finland and Iceland to extend comparisons across countries.

### Statements and Declarations

#### Data availability statement

The data presented in this study were extracted from previously published studies, which are cited in the reference list. No new primary data were generated.

## Acknowledgments

We thank information specialist Susanne Gustafsson at the Swedish Medical Products Agency for her work with the literature search strategy and the literature searches.

## Author contributions

All authors contributed to the conception and/or design of the study. MB, MT, and PT contributed to study selection, data extraction, and quality assessment. MB created the tables and drafted the manuscript. MT created the figures. RL provided supervision throughout the study. All authors interpreted the data. All authors critically revised the manuscript for intellectual content. All authors reviewed and approved the manuscript for submission.

## Disclosures

The authors declare no conflicts of interest.

## Funding

This work was funded by the Government Offices of Sweden, Ministry of Social Affairs (S2024/02156). The funder had no role in the design, data collection, data analysis, and reporting of this study.

## Notes

### Competing Interest Statement

The authors have declared no competing interest.

### Clinical Protocols

https://www.crd.york.ac.uk/PROSPERO/view/CRD420250651803

