## Supplemental file 1 for "Age-sex specific prevalence and incidence of antidepressant prescribing in the Nordic countries: a systematic review"

### Literature search strategy: SSRI and prevalence & incidence

Information specialist: Susanne Gustafsson

**Title: SSRI and prevalence & incidence**

**Database: Pubmed (NLM)**

**Date: 2025-04-04**

| | Search terms | Number of hits |
|---|---|---|
| 1. | "Antidepressive Agents"[Mesh] or "Psychotropic Drugs"[Mesh] or ("anti depress*"[Title/Abstract] or antidepress*[Title/Abstract] or psychotropic*[Title/Abstract] or ssri*[Title/Abstract] or snri*[Title/Abstract] or ssnri*[Title/Abstract]) or "Selective Serotonin Reuptake Inhibitors"[Mesh] or "serotonin inhibitor"[tiab:~3] or "serotonin inhibitors"[tiab:~3] or "noradrenalin inhibitor"[tiab:~3] or "noradrenalin inhibitors"[tiab:~3] or "norepinephrine inhibitor"[tiab:~3] or "norepinephrine inhibitors"[tiab:~3] or "tetracyclic antidepressive"[tiab:~3] or "tetracyclic antidepressant"[tiab:~3] or "tetracyclic antidepressants"[tiab:~3] or "tricyclic antidepressive"[tiab:~3] or "tricyclic antidepressant"[tiab:~3] or "tricyclic antidepressants"[tiab:~3] | 248,163 |
| 2. | "Epidemiology"[Majr] or "Pharmacoepidemiology"[Mesh] or "Incidence"[Mesh] or "Prevalence"[Mesh] or "Drug Utilization"[Majr:NoExp] or epidemiolog*[Title] or incidence*[Title/Abstract] or prevalence[Title/Abstract] or occurrence*[Title] or "point estimate"[Title] or prevalent[Title] or rate[Title] or trend*[Title] or "use of"[Title/Abstract] or "utilization of"[Title] | 4,452,529 |
| 3. | "Scandinavian and Nordic Countries"[Mesh] or "Nordic countr*"[Title/Abstract] or Scandinavia*[Title/Abstract] or Denmark*[Title/Abstract] or Danish[Title/Abstract] or Greenland*[Title/Abstract] or "Faroe Islands"[Title/Abstract] or Finland*[Title/Abstract] or Finnish[Title/Abstract] or Iceland*[Title/Abstract] or Norway*[Title/Abstract] or Norwegian*[Title/Abstract] or Sweden*[Title/Abstract] or Swedish[Title/Abstract] or "countries regions"[ti:~2] or "european countries"[ti:~2] or "europe countries"[ti:~2] or "western countries"[ti:~2] or "western countries"[ti:~2] | 351,953 |
| 4. | 1 and 2 and 3 | 2,183 |
| 5. | (animal[Title/Abstract] or animals[Title/Abstract] or baboon[Title/Abstract] or bird[Title/Abstract] or bovine[Title/Abstract] or canine[Title/Abstract] or cat[Title/Abstract] or cats[Title/Abstract] or cattle[Title/Abstract] or chicken[Title/Abstract] or chickens[Title/Abstract] or cow[Title/Abstract] or cows[Title/Abstract] or dog[Title/Abstract] or dogs[Title/Abstract] or duck[Title/Abstract] or feline[Title/Abstract] or fish[Title/Abstract] or goose[Title/Abstract] or geese[Title/Abstract] or macaque[Title/Abstract] or mouse[Title/Abstract] or mice[Title/Abstract] or murine[Title/Abstract] or nonhuman[Title/Abstract] or ovine[Title/Abstract] or pig[Title/Abstract] or pigs[Title/Abstract] or porcine[Title/Abstract] or primate[Title/Abstract] or rabbit[Title/Abstract] or rabbits[Title/Abstract] or rat[Title/Abstract] or rats[Title/Abstract] or rodent[Title/Abstract] or sheep[Title/Abstract] or swine[Title/Abstract] or zebrafish[Title/Abstract]) | 5,363,446 |
| 6. | 4 not 5 | 2,166 |
| 7. | Filters: Danish, English, Icelandic, Norwegian, Swedish from 2000 - 2025 | **1,884** |

#### Field Tags

- [Mesh] = exploded Mesh term
- [Mesh:noexp] = non exploded Mesh term
- [majr] = Mesh major topic
- [tiab] = title and abstract
- [ot]= other term
- [tw] = text Word
- = truncation of word for alternate endings
- " " = citation marks; searches for exact phrase
- "term term"[Title/Abstract:~2] = proximity operator

**Title: SSRI and prevalence & incidence**

**Database: Embase (Elsevier)**

**Date: 2025-04-04**

| | Search terms | Number of hits |
|---|---|---|
| 1. | 'antidepressant agent'/de or ('anti depress*' or antidepress* or psychotropic* or ssri*):ti,ab,kw or 'noradrenalin uptake inhibitor'/de or ((noradrenalin* or norepinephrin*) near/3 inhibitor*):ti,ab,kw or 'serotonin uptake inhibitor'/de or (serotonin near/3 inhibitor*):ti,ab,kw or 'serotonin noradrenalin reuptake inhibitor'/de or (snri* or ssnri*):ti,ab,kw or 'tetracyclic antidepressant agent'/de or (tetracyclic near/3 antidepress*):ti,ab,kw or 'tricyclic antidepressant agent'/de or (tricyclic near/3 antidepress*):ti,ab,kw | 262,899 |
| 2. | 'prevalence'/de or 'incidence'/exp or 'pharmacoepidemiology'/de or 'drug utilization'/mj or (incidence* or prevalence):ti,ab,kw or (epidemiolog* or occurrence or pattern* or 'point estimate' or prevalent or rate or trend* or 'use of' or 'utilization of'):ti,kw | 4,468,139 |
| 3. | 'Scandinavia'/exp or ('Nordic countr*' or Scandinavia* or Denmark* or Danish or Greenland* or 'Faroe Islands' or Finland* or Finnish or Iceland* or Norway* or Norwegian* or Sweden* or Swedish):ti,ab,kw or ((europe* or western) near/2 countries):ti or 'countries and regions':ti | 415,074 |
| 4. | 1 and 2 and 3 | 2,149 |
| 5. | ('animal'/exp or 'adult animal'/de or 'animal cell'/de or 'animal experiment'/exp or 'animal model'/exp or 'animal tissue'/exp or 'nonhuman'/de) NOT ('human'/exp or 'human experiment'/exp) | 8,264,609 |
| 6. | 4 not 5 | 2,145 |
| 7. | 6 not ([conference abstract]/lim or [conference paper]/lim or [conference review]/lim or [data papers]/lim or [editorial]/lim or [short survey]/lim) | 1,728 |
| 8. | 7 and [2000-2025]/py | **1,625** |

#### Field Tags

- /exp = exploded Emtree term
- /de = non exploded Emtree term
- /mj = major topic
- :ti,ab,kw = title, abstract and author keywords
- = truncation of word for alternate endings
- ' ' = single citation marks; searches for exact phrase
- NEAR/X = within X words, regardless of order
- NEXT/X = terms appear next to each other in the specified order

**Title: SSRI and prevalence & incidence**

**Database: Web of Science (Clarivate)**

**Date: 2025-04-04**

| | Search terms | Number of hits |
|---|---|---|
| 1. | TI=("anti depress*" or antidepress* or psychotropic* or ssri* or snri* or ssnri*) or AK=("anti depress*" or antidepress* or psychotropic* or ssri* or snri* or ssnri*) or AB=("anti depress*" or antidepress* or psychotropic* or ssri* or snri* or ssnri*) or TI=((noradrenalin* or norepinephrin* or serotonin) near/3 inhibitor*) or AK=((noradrenalin* or norepinephrin* or serotonin) near/3 inhibitor*) or AB=((noradrenalin* or norepinephrin* or serotonin) near/3 inhibitor*) or TI=((tetracyclic or tricyclic) near/3 antidepress*) or AK=((tetracyclic or tricyclic) near/3 antidepress*) or AB=((tetracyclic or tricyclic) near/3 antidepress*) | 130,167 |
| 2. | TI=(incidence* or prevalence) or AB=(incidence* or prevalence) or AK=(incidence* or prevalence) or TI=(epidemiolog* or occurrence or pattern* or pharmacoepidemiology or "point estimate" or prevalent or rate or trend* or "use of" or "utilization of") or AK=(epidemiolog* or occurrence or pattern* or pharmacoepidemiology or "point estimate" or prevalent or rate or trend* or "use of" or "utilization of") | 4,522,711 |
| 3. | TI=("Nordic countr*" or Scandinavia* or Denmark* or Danish or Greenland* or "Faroe Islands" or Finland* or Finnish or Iceland* or Norway* or Norwegian* or Sweden* or Swedish) or AB=("Nordic countr*" or Scandinavia* or Denmark* or Danish or Greenland* or "Faroe Islands" or Finland* or Finnish or Iceland* or Norway* or Norwegian* or Sweden* or Swedish) or AK=("Nordic countr*" or Scandinavia* or Denmark* or Danish or Greenland* or "Faroe Islands" or Finland* or Finnish or Iceland* or Norway* or Norwegian* or Sweden* or Swedish) or TI=((europe* or western) near/2 countries) or TI="countries and regions" | 523,135 |
| 4. | 1 and 2 and 3 | 1,094 |
| 5. | TS=(animal* or avian* or baboon* or bird* or bovine or canine or cat or cats or cattle* or chicken or chickens or cow or cows or dog or dogs or duck or feline or fish* or goose or geese or macaque* or mouse or mice or murine or nonhuman* or ovine or pig or pigs or porcine or primate* or rabbit or rabbits or rat or rats or rodent* or sheep or swine or veterinar* or zebrafish*) | 7,942,663 |
| 6. | 4 not 5 | 1,081 |
| 7. | Document types: Article or Early Access or Letter or Note and Languages: English + Scandinavian languages and Publication Years: 2000 - 2025 | **960** |

**Field tags**

- AK=author keywords
- AB = abstract
- TI = title
- Topic = title, abstract, author keywords and keywords plus
- * = truncation of word for alternate endings
- " " = citation marks; searches for exact phrase
- NEAR/X = within X words, regardless of order

Before deduplication: 4,469
After deduplication: 2,595
