## Supplemental table 1 for "Age-sex specific prevalence and incidence of antidepressant prescribing in the Nordic countries: a systematic review"

| **Supplemental Table 1.** Excluded studies and reasons for exclusion | | |
| --- | --- | --- |
| **Reason for exclusion** | **Number of studies** | **Study** |
| Wrong outcome | 11 | Bramness, J. G., Hausken, A. M., Sakshaug, S., Skurtveit, S., & Rønning, M. (2005). Prescription of selective serotonin reuptake inhibitors 1990-2004. Tidsskrift for den Norske Laegeforening: Tidsskrift for Praktisk Medicin, ny Raekke, 125(18), 2470-2473.  Eek, E., van Driel, M., Falk, M., Hollingworth, S. A., & Merlo, G. (2021). Antidepressant use in Australia and Sweden—A cross‐country comparison. Pharmacoepidemiology and drug safety, 30(4), 409-417.  Forns, J., Pottegård, A., Reinders, T., Poblador-Plou, B., Morros, R., Brandt, L., ... & Reutfors, J. (2019). Antidepressant use in Denmark, Germany, Spain, and Sweden between 2009 and 2014: Incidence and comorbidities of antidepressant initiators. Journal of affective disorders, 249, 242-252.  Kildegaard, H., Wesselhoeft, R., Lund, L. C., & Bliddal, M. (2024). Post-pandemic trends in psychotropic medication use in Danish children, adolescents, and young adults. Acta Psychiatrica Scandinavica, 150(3), 174-177.  Pasman, J. A., Meijsen, J. J., Haram, M., Kowalec, K., Harder, A., Xiong, Y., ... & Lu, Y. (2023). Epidemiological overview of major depressive disorder in Scandinavia using nationwide registers. *The Lancet Regional Health–Europe*, *29*.  Pedersen, E., Tripodi, E., Aakjær, M., Li, H., Cantarutti, A., Nyberg, F., ... & Nordeng, H. (2024). Drug utilisation in children and adolescents before and after the start of the COVID‐19 pandemic: interrupted time‐series analyses in three European countries. Paediatric and Perinatal Epidemiology, 38(6), 450-460.  Rosland, H. G., Wergeland, G. J., & Holst, L. (2025). Off-label use of psychotropic drugs in youth. BMC psychiatry, 25(1), 739.  Samardžić, J., Simović, F., Sekanić, K., & Branković, M. (2025, May). Five-Year Trends in SSRI Consumption: A Precision Medicine Approach to Comparative Analysis Between Serbia and European Countries. In Healthcare (Vol. 13, No. 10, p. 1174). MDPI.  Seljeflot, L. M., Blix, H. S., Andersson, Y., & Hynnekleiv, T. (2025). Use of psychotropic medications in the specialist health service for mental health care and substance use disorders in 2012–23. Tidsskrift for Den norske legeforening.  Tomasson, K., Tomasson, H., Zoega, T., Sigfusson, E., & Helgason, T. (2007). Epidemiology of psychotropic medication use: comparison of sales, prescriptions and survey data in Iceland. Nordic journal of psychiatry, 61(6), 471-478.  Wolfschlag, M., Grudet, C., & Håkansson, A. (2021). Impact of the COVID-19 pandemic on the general mental health in Sweden: no observed changes in the dispensed amount of common psychotropic medications in the region of Scania. *Frontiers in psychiatry*, *12*, 731297. |
| Wrong study objective | 5 | Bramness, J. G., Engeland, A., & Furu, K. (2007). Use of antidepressants among children and adolescents--did the warnings lead to fewer prescriptions?. *Tidsskrift for den Norske laegeforening: tidsskrift for praktisk medicin, ny raekke*, *127*(20), 2653-2655.  Geest, A., Bonnesen, B., Jordan, A., Tønnesen, L., Rømer, V., Ulrik, C. S., ... & Jensen, J. U. S. (2025). The impact of social distancing on mental health during the COVID-19 pandemic: a nationwide study of 4.6 million Danish adults. *European Psychiatry*, *68*(1), e30.  Gøtzsche, P. C. (2020). Long-term use of antipsychotics and antidepressants is not evidence-based. *International journal of risk & safety in medicine*, *31*(1), 37-42.  Nielsen, E. S., Rasmussen, L., Hellfritzsch, M., Thomsen, P. H., Nørgaard, M., & Laursen, T. (2017). Trends in Off‐Label Prescribing of Sedatives, Hypnotics and Antidepressants among Children and Adolescents–A Danish, Nationwide Register‐Based Study. Basic & Clinical Pharmacology & Toxicology, 120(4), 360-367.  von Heideken Wågert, P., Gustavsson, J. M., Lundin-Olsson, L., Kallin, K., Nygren, B., Lundman, B., ... & Gustafson, Y. (2006). Health status in the oldest old: Age and sex differences in the Umeå 85+ Study. *Aging clinical and experimental research*, *18*(2), 116-126. |
| Wrong population | 4 | Hanson, L. L. M., Madsen, I. E., Westerlund, H., Theorell, T., Burr, H., & Rugulies, R. (2013). Antidepressant use and associations with psychosocial work characteristics. A comparative study of Swedish and Danish gainfully employed. Journal of affective disorders, 149(1-3), 38-45.  Hentilä, E., Tiihonen, M., Taipale, H., Hartikainen, S., & Tolppanen, A. M. (2021). Incidence of antidepressant use among community dwellers with and without Parkinson’s disease–a nationwide cohort study. *BMC geriatrics*, *21*(1), 202.  Puranen, A., Taipale, H., Koponen, M., Tanskanen, A., Tolppanen, A. M., Tiihonen, J., & Hartikainen, S. (2017). Incidence of antidepressant use in community‐dwelling persons with and without Alzheimer's disease: 13‐year follow‐up. *International journal of geriatric psychiatry*, *32*(1), 94-101.  Taipale, H., Koponen, M., Tanskanen, A., Tolppanen, A. M., Tiihonen, J., & Hartikainen, S. (2016). Drug use in persons with and without Alzheimer's disease aged 90 years or more. *Age and ageing*, *45*(6), 900-904. |
| Wrong drug | 2 | Andersen‐Ranberg, K., Schroll, M., & Jeune, B. (2001). Healthy centenarians do not exist, but autonomous centenarians do: a population‐based study of morbidity among Danish centenarians. *Journal of the American Geriatrics Society*, *49*(7), 900-908.  Dahlén, E., & Kimland, E. E. (2024). Considerable paediatric drug dispensing–A nationwide study of more than 2 million Swedish children. *Acta Paediatrica*, *113*(9), 2147-2154. |
| Wrong publication type | 1 | Svaleryd, H., Björkegren, E., & Vlachos, J. (2021). The impact of the COVID-19 school closure on adolescents’ use of mental healthcare services in Sweden. MedRxiv, 2021-12. |
| Data could neither be extracted from the article nor provided by author upon request | 1 | Hartikainen, S., & Klaukka, T. (2004). Use of psychotropics is high among very old people. *European Journal of Clinical Pharmacology*, *59*(11), 849-850. |
