## Supplemental table 2 for "Age-sex specific prevalence and incidence of antidepressant prescribing in the Nordic countries: a systematic review"

| **Supplemental table 2.** Study quality in individual studies according to the Joanna Briggs Institute Critical Appraisal Checklist for Studies Reporting Prevalence Data. | | | | | | | | | | |
| --- | --- | --- | --- | --- | --- | --- | --- | --- | --- | --- |
| **Author (year)** | **Sample frame appropriate to address target population?** | **Participants sampled appropriately?** | **Sample size adequate?** | **Subjects and setting described in detail?** | **Data analysis conducted with sufficient coverage of the identified sample?** | **Valid methods for identification of the condition?** | **Condition measured in standard, reliable way for all participants?** | **Appropriate statistical analysis?** | **Adequate response rate? If not, low response rate managed appropriately?** | **Total score (0-9)** |
| Abbing-Karahagopian 2014 | Yes | Yes | Yes | No | Yes | Yes | Yes | Yes | Yes | 8 |
| Autti-Rämö 2011 | Yes | Yes | Yes | No | Yes | Yes | Yes | No | Yes | 7 |
| Bachmann 2016 | Yes | Yes | Yes | No | Yes | Yes | Yes | No | Yes | 7 |
| Bojanić 2024 | Yes | Yes | Yes | No | Yes | Yes | Yes | No | Yes | 7 |
| Corneliusson 2024 | Yes | Yes | Unclear | Yes | Unclear | Yes | Yes | No | Unclear | 5 |
| Forslund 2020 | Yes | Yes | Yes | No | Yes | Yes | Yes | No | Yes | 7 |
| Foulon 2010 | Yes | Yes | Yes | No | Yes | Yes | Yes | No | Yes | 7 |
| Gómez-Lumbreras 2021 | Yes | Yes | Yes | No | Yes | Yes | Yes | No | Yes | 7 |
| Hansen 2007 | Yes | Yes | Yes | No | Yes | Yes | Yes | No | Yes | 7 |
| Hartz 2016a | Yes | Yes | Yes | No | Yes | Yes | Yes | No | Yes | 7 |
| Hartz 2016b | Yes | Yes | Yes | No | Yes | Yes | Yes | No | Yes | 7 |
| Ingemann 2021 | Yes | Yes | Yes | No | Yes | Yes | Yes | Yes | Yes | 8 |
| Ishtiak-Ahmed 2023 | Yes | Yes | Yes | No | Yes | Yes | Yes | No | Yes | 7 |
| Kjosavik 2011 | Yes | Yes | Yes | No | Yes | Yes | Yes | No | Yes | 7 |
| Kjosavik 2009 | Yes | Yes | Yes | No | Yes | Yes | Yes | No | Yes | 7 |
| Lagerberg 2019 | Yes | Yes | Yes | No | Yes | Yes | Yes | No | Yes | 7 |
| Lien 2023 | Yes | Yes | Yes | No | Yes | Yes | Yes | No | Yes | 7 |
| Loikas 2013 | Yes | Yes | Yes | No | Yes | Yes | Yes | No | Yes | 7 |
| Pottegård 2014 | Yes | Yes | Yes | No | Yes | Yes | Yes | No | Yes | 7 |
| Rasmussen 2024 | Yes | Yes | Yes | No | Yes | Yes | Yes | No | Yes | 7 |
| Saastamoinen 2012 | Yes | Yes | Yes | No | Yes | Yes | Yes | No | Yes | 7 |
| Sihvo 2008 | Yes | Yes | Yes | Yes | Yes | Yes | Yes | No | Yes | 8 |
| Sihvo 2010 | Yes | Yes | Yes | Yes | Yes | Yes | Yes | No | Yes | 8 |
| Skovlund 2017 | Yes | Yes | Yes | No | Yes | Yes | Yes | No | Yes | 7 |
| Steffenak 2012 | Yes | Yes | Yes | No | Yes | Yes | Yes | No | Yes | 7 |
| Steinhausen 2014 | Yes | Yes | Yes | No | Yes | Yes | Yes | No | Yes | 7 |
| Wastesson 2012 | Yes | Yes | Yes | No | Yes | Yes | Yes | No | Yes | 7 |
| Wesselhoeft 2020 | Yes | Yes | Yes | No | Yes | Yes | Yes | No | Yes | 7 |
| Zito 2006 | Yes | Yes | Yes | No | Yes | Yes | Yes | No | Yes | 7 |
| Zoëga 2009 | Yes | Yes | Yes | No | Yes | Yes | Yes | No | Yes | 7 |
