## Supplemental table 3 for "Age-sex specific prevalence and incidence of antidepressant prescribing in the Nordic countries: a systematic review"

| **Supplemental table 3.** Results in studies on prevalence of antidepressant prescription. | | | | | |
| --- | --- | --- | --- | --- | --- |
| **Study** | **Country** | **Year** | **Sex** | **Ages** | **Prevalence per 1000** |
| Zito 2006^a^ [] | Denmark | 2000 | Combined | 0-19 | 1.75 |
|  |  |  | Combined | 0-4 | 0.03 |
|  |  |  | Combined | 5-9 | 0.36 |
|  |  |  | Combined | 10-14 | 1.01 |
|  |  |  | Combined | 15-19 | 6.21 |
|  |  |  | Male | 0-19 | 1.3 |
|  |  |  | Male | 0-4 | - |
|  |  |  | Male | 5-9 | 0.45 |
|  |  |  | Male | 10-14 | 1.17 |
|  |  |  | Male | 15-19 | 3.98 |
|  |  |  | Female | 0-19 | 2.23 |
|  |  |  | Female | 0-4 | 0.07 |
|  |  |  | Female | 5-9 | 0.27 |
|  |  |  | Female | 10-14 | 0.85 |
|  |  |  | Female | 15-19 | 8.53 |
| Steinhausen 2014^b^ [] | Denmark | 1996 | Combined | 0-17 | 0.28 |
|  |  | 1997 | Combined | 0-17 | 0.46 |
|  |  | 1998 | Combined | 0-17 | 0.61 |
|  |  | 1999 | Combined | 0-17 | 0.78 |
|  |  | 2000 | Combined | 0-17 | 0.96 |
|  |  | 2001 | Combined | 0-17 | 1.29 |
|  |  | 2002 | Combined | 0-17 | 1.57 |
|  |  | 2003 | Combined | 0-17 | 2 |
|  |  | 2004 | Combined | 0-17 | 2.01 |
|  |  | 2005 | Combined | 0-17 | 2.15 |
|  |  | 2006 | Combined | 0-17 | 2.35 |
|  |  | 2007 | Combined | 0-17 | 2.53 |
|  |  | 2008 | Combined | 0-17 | 2.69 |
|  |  | 2009 | Combined | 0-17 | 2.74 |
|  |  | 2010 | Combined | 0-17 | 2.67 |
| Skovlund 2017^a^ [] | Denmark | 2000 | Male | 10-14 | 1.85 |
|  |  | 2001 | Male | 10-14 | 2.16 |
|  |  | 2002 | Male | 10-14 | 2.59 |
|  |  | 2003 | Male | 10-14 | 3.01 |
|  |  | 2004 | Male | 10-14 | 2.95 |
|  |  | 2005 | Male | 10-14 | 2.99 |
|  |  | 2006 | Male | 10-14 | 3.26 |
|  |  | 2007 | Male | 10-14 | 3.54 |
|  |  | 2008 | Male | 10-14 | 3.87 |
|  |  | 2009 | Male | 10-14 | 4.13 |
|  |  | 2010 | Male | 10-14 | 4.44 |
|  |  | 2011 | Male | 10-14 | 4.36 |
|  |  | 2012 | Male | 10-14 | 4.35 |
|  |  | 2013 | Male | 10-14 | 4.02 |
|  |  | 2000 | Female | 10-14 | 1.67 |
|  |  | 2001 | Female | 10-14 | 2.39 |
|  |  | 2002 | Female | 10-14 | 2.89 |
|  |  | 2003 | Female | 10-14 | 3.65 |
|  |  | 2004 | Female | 10-14 | 3.69 |
|  |  | 2005 | Female | 10-14 | 4.2 |
|  |  | 2006 | Female | 10-14 | 4.5 |
|  |  | 2007 | Female | 10-14 | 4.7 |
|  |  | 2008 | Female | 10-14 | 4.95 |
|  |  | 2009 | Female | 10-14 | 5.25 |
|  |  | 2010 | Female | 10-14 | 6.07 |
|  |  | 2011 | Female | 10-14 | 5.12 |
|  |  | 2012 | Female | 10-14 | 5.12 |
|  |  | 2013 | Female | 10-14 | 4.79 |
|  |  | 2000 | Male | 15-19 | 5.5 |
|  |  | 2001 | Male | 15-19 | 7.1 |
|  |  | 2002 | Male | 15-19 | 8.6 |
|  |  | 2003 | Male | 15-19 | 10.3 |
|  |  | 2004 | Male | 15-19 | 10.6 |
|  |  | 2005 | Male | 15-19 | 12.5 |
|  |  | 2006 | Male | 15-19 | 14.3 |
|  |  | 2007 | Male | 15-19 | 15.6 |
|  |  | 2008 | Male | 15-19 | 16.6 |
|  |  | 2009 | Male | 15-19 | 18.7 |
|  |  | 2010 | Male | 15-19 | 21 |
|  |  | 2011 | Male | 15-19 | 19.8 |
|  |  | 2012 | Male | 15-19 | 17.8 |
|  |  | 2013 | Male | 15-19 | 15.7 |
|  |  | 2000 | Female | 15-19 | 13.1 |
|  |  | 2001 | Female | 15-19 | 17.4 |
|  |  | 2002 | Female | 15-19 | 21.2 |
|  |  | 2003 | Female | 15-19 | 26.2 |
|  |  | 2004 | Female | 15-19 | 27.1 |
|  |  | 2005 | Female | 15-19 | 30 |
|  |  | 2006 | Female | 15-19 | 33.9 |
|  |  | 2007 | Female | 15-19 | 37.5 |
|  |  | 2008 | Female | 15-19 | 41.1 |
|  |  | 2009 | Female | 15-19 | 44.4 |
|  |  | 2010 | Female | 15-19 | 50 |
|  |  | 2011 | Female | 15-19 | 46.9 |
|  |  | 2012 | Female | 15-19 | 43.8 |
|  |  | 2013 | Female | 15-19 | 38.5 |
| Bojanić 2024^a^ [] | Denmark | 2006 | Combined | All | 72.8 |
|  |  |  | Combined | 5-19 | 8.8 |
|  |  |  | Combined | 20-65 | 80.5 |
|  |  |  | Combined | 65 and older | 144 |
|  |  |  | Male | 15-19 | 13.4 |
|  |  |  | Male | 20-44 | 46.4 |
|  |  |  | Male | 45-64 | 71.9 |
|  |  |  | Male | 65-74 | 89.7 |
|  |  |  | Male | 75 and older | 170.1 |
|  |  |  | Female | 15-19 | 32.2 |
|  |  |  | Female | 20-44 | 81.3 |
|  |  |  | Female | 45-64 | 121.5 |
|  |  |  | Female | 65-74 | 130.7 |
|  |  |  | Female | 75 and older | 246.8 |
|  |  | 2009 | Combined | 5 and older | 80.2 |
|  |  |  | Combined | 5-19 | 12.1 |
|  |  |  | Combined | 20-65 | 90.7 |
|  |  |  | Combined | 65 and older | 150.4 |
|  |  |  | Male | 15-19 | 17.6 |
|  |  |  | Male | 20-44 | 53.4 |
|  |  |  | Male | 45-64 | 78.9 |
|  |  |  | Male | 65-74 | 94.9 |
|  |  |  | Male | 75 and older | 177.2 |
|  |  |  | Female | 15-19 | 42.1 |
|  |  |  | Female | 20-44 | 95 |
|  |  |  | Female | 45-64 | 131.1 |
|  |  |  | Female | 65-74 | 134.8 |
|  |  |  | Female | 75 and older | 255.4 |
|  |  | 2012 | Combined | 5 and older | 82.2 |
|  |  |  | Combined | 5-19 | 12.3 |
|  |  |  | Combined | 20-65 | 94 |
|  |  |  | Combined | 65 and older | 140.3 |
|  |  |  | Male | 15-19 | 16.6 |
|  |  |  | Male | 20-44 | 56.4 |
|  |  |  | Male | 45-64 | 81.2 |
|  |  |  | Male | 65-74 | 92.5 |
|  |  |  | Male | 75 and older | 173.5 |
|  |  |  | Female | 15-19 | 40.8 |
|  |  |  | Female | 20-44 | 99.4 |
|  |  |  | Female | 45-64 | 133.9 |
|  |  |  | Female | 65-74 | 129.7 |
|  |  |  | Female | 75 and older | 252.3 |
|  |  | 2015 | Combined | 5 and older | 74.4 |
|  |  |  | Combined | 5-19 | 8.8 |
|  |  |  | Combined | 20-65 | 83 |
|  |  |  | Combined | 65 and older | 129.2 |
|  |  |  | Male | 15-19 | 11.1 |
|  |  |  | Male | 20-44 | 47.9 |
|  |  |  | Male | 45-64 | 74.4 |
|  |  |  | Male | 65-74 | 85.4 |
|  |  |  | Male | 75 and older | 156.7 |
|  |  |  | Female | 15-19 | 28.6 |
|  |  |  | Female | 20-44 | 84.3 |
|  |  |  | Female | 45-64 | 124.6 |
|  |  |  | Female | 65-74 | 118.6 |
|  |  |  | Female | 75 and older | 232.1 |
|  |  | 2018 | Combined | 5 and older | 72.2 |
|  |  |  | Combined | 5-19 | 8.4 |
|  |  |  | Combined | 20-65 | 79.3 |
|  |  |  | Combined | 65 and older | 125.6 |
|  |  |  | Male | 15-19 | 12.1 |
|  |  |  | Male | 20-44 | 45.6 |
|  |  |  | Male | 45-64 | 72.5 |
|  |  |  | Male | 65-74 | 84.1 |
|  |  |  | Male | 75 and older | 145.2 |
|  |  |  | Female | 15-19 | 26.6 |
|  |  |  | Female | 20-44 | 78.9 |
|  |  |  | Female | 45-64 | 121.5 |
|  |  |  | Female | 65-74 | 116.4 |
|  |  |  | Female | 75 and older | 218.7 |
|  |  | 2021 | Combined | 5 and older | 76.6 |
|  |  |  | Combined | 5-19 | 11 |
|  |  |  | Combined | 20-65 | 84.6 |
|  |  |  | Combined | 65 and older | 126.4 |
|  |  |  | Male | 15-19 | 14.5 |
|  |  |  | Male | 20-44 | 50.3 |
|  |  |  | Male | 45-64 | 74.2 |
|  |  |  | Male | 65-74 | 86.4 |
|  |  |  | Male | 75 and older | 142.6 |
|  |  |  | Female | 15-19 | 35.8 |
|  |  |  | Female | 20-44 | 88.2 |
|  |  |  | Female | 45-64 | 125.6 |
|  |  |  | Female | 65-74 | 120.9 |
|  |  |  | Female | 75 and older | 213.6 |
| Gómez-Lumbreras 2021^a^ [] | Denmark | 2008 | Combined | 0-17 | 4.49 |
|  |  |  | Male | 0-17 | 3.08 |
|  |  |  | Female | 0-17 | 5.98 |
|  |  | 2009 | Combined | 0-17 | 4.86 |
|  |  |  | Male | 0-17 | 3.37 |
|  |  |  | Female | 0-17 | 6.42 |
|  |  | 2010 | Combined | 0-17 | 5.43 |
|  |  |  | Male | 0-17 | 3.68 |
|  |  |  | Female | 0-17 | 7.27 |
|  |  | 2011 | Combined | 0-17 | 5.06 |
|  |  |  | Male | 0-17 | 3.63 |
|  |  |  | Female | 0-17 | 6.55 |
|  |  | 2012 | Combined | 0-17 | 4.72 |
|  |  |  | Male | 0-17 | 3.33 |
|  |  |  | Female | 0-17 | 6.17 |
|  |  | 2013 | Combined | 0-17 | 4.17 |
|  |  |  | Male | 0-17 | 2.95 |
|  |  |  | Female | 0-17 | 5.46 |
|  |  | 2014 | Combined | 0-17 | 3.65 |
|  |  |  | Male | 0-17 | 2.55 |
|  |  |  | Female | 0-17 | 4.81 |
|  |  | 2015 | Combined | 0-17 | 3.5 |
|  |  |  | Male | 0-17 | 2.44 |
|  |  |  | Female | 0-17 | 4.62 |
|  |  | 2016 | Combined | 0-17 | 3.33 |
|  |  |  | Male | 0-17 | 2.38 |
|  |  |  | Female | 0-17 | 4.33 |
|  |  | 2017 | Combined | 0-17 | 3.1 |
|  |  |  | Male | 0-17 | 2.3 |
|  |  |  | Male | 0-4 | 0 |
|  |  |  | Male | 5-9 | 0.24 |
|  |  |  | Male | 10-14 | 2.84 |
|  |  |  | Male | 15-17 | 7.9 |
|  |  |  | Female | 0-17 | 4.11 |
|  |  |  | Female | 0-4 | 0 |
|  |  |  | Female | 5-9 | 0.09 |
|  |  |  | Female | 10-14 | 3.02 |
|  |  |  | Female | 15-17 | 17.95 |
| Wesselhoeft 2020^a^ [] | Denmark | 2007 | Combined | 5-19 | 9.27 |
|  |  | 2008 | Combined | 5-19 | 10.23 |
|  |  | 2009 | Combined | 5-19 | 11.32 |
|  |  | 2010 | Combined | 5-19 | 12.86 |
|  |  | 2011 | Combined | 5-19 | 12.26 |
|  |  | 2012 | Combined | 5-19 | 11.45 |
|  |  | 2013 | Combined | 5-19 | 10.17 |
|  |  | 2014 | Combined | 5-19 | 8.81 |
|  |  | 2015 | Combined | 5-19 | 8.12 |
|  |  | 2016 | Combined | 5-19 | 7.67 |
|  |  | 2017 | Combined | 5-19 | 7.52 |
|  |  | 2007 | Female | 5-9 | 0.26 |
|  |  | 2007 | Male | 5-9 | 0.57 |
|  |  | 2007 | Female | 10-14 | 3.80 |
|  |  | 2007 | Male | 10-14 | 3.21 |
|  |  | 2007 | Female | 15-19 | 35.52 |
|  |  | 2007 | Male | 15-19 | 14.65 |
|  |  | 2008 | Female | 5-9 | 0.23 |
|  |  | 2008 | Male | 5-9 | 0.72 |
|  |  | 2008 | Female | 10-14 | 3.96 |
|  |  | 2008 | Male | 10-14 | 3.48 |
|  |  | 2008 | Female | 15-19 | 38.78 |
|  |  | 2008 | Male | 15-19 | 15.44 |
|  |  | 2009 | Female | 5-9 | 0.43 |
|  |  | 2009 | Male | 5-9 | 0.62 |
|  |  | 2009 | Female | 10-14 | 4.14 |
|  |  | 2009 | Male | 10-14 | 3.74 |
|  |  | 2009 | Female | 15-19 | 41.74 |
|  |  | 2009 | Male | 15-19 | 17.47 |
|  |  | 2010 | Female | 5-9 | 0.40 |
|  |  | 2010 | Male | 5-9 | 0.69 |
|  |  | 2010 | Female | 10-14 | 4.80 |
|  |  | 2010 | Male | 10-14 | 4.05 |
|  |  | 2010 | Female | 15-19 | 46.90 |
|  |  | 2010 | Male | 15-19 | 19.27 |
|  |  | 2011 | Female | 5-9 | 0.44 |
|  |  | 2011 | Male | 5-9 | 0.60 |
|  |  | 2011 | Female | 10-14 | 4.23 |
|  |  | 2011 | Male | 10-14 | 4.07 |
|  |  | 2011 | Female | 15-19 | 43.88 |
|  |  | 2011 | Male | 15-19 | 18.42 |
|  |  | 2012 | Female | 5-9 | 0.35 |
|  |  | 2012 | Male | 5-9 | 0.52 |
|  |  | 2012 | Female | 10-14 | 4.26 |
|  |  | 2012 | Male | 10-14 | 3.99 |
|  |  | 2012 | Female | 15-19 | 40.89 |
|  |  | 2012 | Male | 15-19 | 16.61 |
|  |  | 2013 | Female | 5-9 | 0.27 |
|  |  | 2013 | Male | 5-9 | 0.36 |
|  |  | 2013 | Female | 10-14 | 4.01 |
|  |  | 2013 | Male | 10-14 | 3.81 |
|  |  | 2013 | Female | 15-19 | 36.13 |
|  |  | 2013 | Male | 15-19 | 14.62 |
|  |  | 2014 | Female | 5-9 | 0.24 |
|  |  | 2014 | Male | 5-9 | 0.37 |
|  |  | 2014 | Female | 10-14 | 3.56 |
|  |  | 2014 | Male | 10-14 | 3.52 |
|  |  | 2014 | Female | 15-19 | 31.48 |
|  |  | 2014 | Male | 15-19 | 12.28 |
|  |  | 2015 | Female | 5-9 | 0.24 |
|  |  | 2015 | Male | 5-9 | 0.37 |
|  |  | 2015 | Female | 10-14 | 3.54 |
|  |  | 2015 | Male | 10-14 | 3.41 |
|  |  | 2015 | Female | 15-19 | 28.91 |
|  |  | 2015 | Male | 15-19 | 11.25 |
|  |  | 2016 | Female | 5-9 | 0.20 |
|  |  | 2016 | Male | 5-9 | 0.33 |
|  |  | 2016 | Female | 10-14 | 3.28 |
|  |  | 2016 | Male | 10-14 | 3.09 |
|  |  | 2016 | Female | 15-19 | 27.19 |
|  |  | 2016 | Male | 15-19 | 11.08 |
|  |  | 2017 | Female | 5-9 | 0.09 |
|  |  | 2017 | Male | 5-9 | 0.24 |
|  |  | 2017 | Female | 10-14 | 3.02 |
|  |  | 2017 | Male | 10-14 | 2.84 |
|  |  | 2017 | Female | 15-19 | 26.96 |
|  |  | 2017 | Male | 15-19 | 11.06 |
| Hansen 2007^a^ [] | Denmark | 2004 | Male | 20-64 | 50 |
|  |  |  | Male | 65-69 | 82 |
|  |  |  | Male | 70-74 | 91 |
|  |  |  | Male | 75-79 | 118 |
|  |  |  | Male | 80-84 | 150 |
|  |  |  | Male | 85 and older | 175 |
|  |  |  | Female | 20-64 | 88 |
|  |  |  | Female | 65-69 | 125 |
|  |  |  | Female | 70-74 | 154 |
|  |  |  | Female | 75-79 | 182 |
|  |  |  | Female | 80-84 | 216 |
|  |  |  | Female | 85-89 | 268 |
|  |  |  | Female | 90 and older | 255 |
| Ingemann 2021^a^ [] | Denmark | 2019 | Combined | 10-89 | 81.50 |
|  |  |  | Combined | 10-19 | 11.62 |
|  |  |  | Combined | 20-29 | 47.83 |
|  |  |  | Combined | 30-39 | 67.59 |
|  |  |  | Combined | 40-49 | 86.91 |
|  |  |  | Combined | 50-59 | 96.46 |
|  |  |  | Combined | 60-69 | 104.99 |
|  |  |  | Combined | 70-79 | 120.81 |
|  |  |  | Combined | 80-89 | 183.34 |
|  |  |  | Male | 10-89 | 59.03 |
|  |  |  | Male | 10-19 | 8.05 |
|  |  |  | Male | 20-29 | 34.84 |
|  |  |  | Male | 30-39 | 50.54 |
|  |  |  | Male | 40-49 | 63.75 |
|  |  |  | Male | 50-59 | 71.88 |
|  |  |  | Male | 60-69 | 80.55 |
|  |  |  | Male | 70-79 | 93.82 |
|  |  |  | Male | 80-89 | 139.73 |
|  |  |  | Female | 10-89 | 103.75 |
|  |  |  | Female | 10-19 | 15.37 |
|  |  |  | Female | 20-29 | 61.37 |
|  |  |  | Female | 30-39 | 85.19 |
|  |  |  | Female | 40-49 | 110.23 |
|  |  |  | Female | 50-59 | 121.29 |
|  |  |  | Female | 60-69 | 128.64 |
|  |  |  | Female | 70-79 | 145.26 |
|  |  |  | Female | 80-89 | 214.45 |
|  | Greenland | 2019 | Combined | 10-89 | 34.54 |
|  |  |  | Combined | 10-19 | 10.67 |
|  |  |  | Combined | 20-29 | 25.65 |
|  |  |  | Combined | 30-39 | 28.16 |
|  |  |  | Combined | 40-49 | 43.40 |
|  |  |  | Combined | 50-59 | 39.55 |
|  |  |  | Combined | 60-69 | 44.16 |
|  |  |  | Combined | 70-79 | 79.37 |
|  |  |  | Combined | 80-89 | 123.81 |
|  |  |  | Male | 10-89 | 20.97 |
|  |  |  | Male | 10-19 | 5.43 |
|  |  |  | Male | 20-29 | 14.97 |
|  |  |  | Male | 30-39 | 18.04 |
|  |  |  | Male | 40-49 | 26.32 |
|  |  |  | Male | 50-59 | 22.76 |
|  |  |  | Male | 60-69 | 29.80 |
|  |  |  | Male | 70-79 | 45.65 |
|  |  |  | Male | 80-89 | 78.34 |
|  |  |  | Female | 10-89 | 49.84 |
|  |  |  | Female | 10-19 | 16.14 |
|  |  |  | Female | 20-29 | 37.01 |
|  |  |  | Female | 30-39 | 38.92 |
|  |  |  | Female | 40-49 | 63.89 |
|  |  |  | Female | 50-59 | 59.42 |
|  |  |  | Female | 60-69 | 62.70 |
|  |  |  | Female | 70-79 | 120.89 |
|  |  |  | Female | 80-89 | 155.84 |
| Abbing-Karahagopian 2014^b^ [] | Denmark | 2008 | Combined | All ages | 61.6 |
| Pottegård 2014^b^ [] | Denmark | 1995 | Combined | 5-17 | 0.1 |
|  |  | 2010 | Combined | 5-17 | 3.3 |
| Bachmann 2016^a^ [] | Denmark | 2005 | Combined | 0-19 | 6.09 |
|  |  |  | Combined | 0-4 | 0.05 |
|  |  |  | Combined | 5-9 | 0.46 |
|  |  |  | Combined | 10-14 | 3.42 |
|  |  |  | Combined | 15-19 | 22.00 |
|  |  |  | Male | 0-19 | 3.96 |
|  |  |  | Female | 0-19 | 8.34 |
|  |  | 2006 | Combined | 0-19 | 6.94 |
|  |  |  | Combined | 0-4 | 0.05 |
|  |  |  | Combined | 5-9 | 0.48 |
|  |  |  | Combined | 10-14 | 3.76 |
|  |  |  | Combined | 15-19 | 24.68 |
|  |  |  | Male | 0-19 | 4.52 |
|  |  |  | Female | 0-19 | 9.49 |
|  |  | 2007 | Combined | 0-19 | 7.82 |
|  |  |  | Combined | 0-4 | 0.06 |
|  |  |  | Combined | 5-9 | 0.47 |
|  |  |  | Combined | 10-14 | 3.88 |
|  |  |  | Combined | 15-19 | 27.69 |
|  |  |  | Male | 0-19 | 5.05 |
|  |  |  | Female | 0-19 | 10.72 |
|  |  | 2008 | Combined | 0-19 | 8.64 |
|  |  |  | Combined | 0-4 | 0.06 |
|  |  |  | Combined | 5-9 | 0.54 |
|  |  |  | Combined | 10-14 | 4.13 |
|  |  |  | Combined | 15-19 | 29.92 |
|  |  |  | Male | 0-19 | 5.50 |
|  |  |  | Female | 0-19 | 11.95 |
|  |  | 2009 | Combined | 0-19 | 9.58 |
|  |  |  | Combined | 0-4 | 0.06 |
|  |  |  | Combined | 5-9 | 0.59 |
|  |  |  | Combined | 10-14 | 4.39 |
|  |  |  | Combined | 15-19 | 32.77 |
|  |  |  | Male | 0-19 | 6.21 |
|  |  |  | Female | 0-19 | 13.12 |
|  |  | 2010 | Combined | 0-19 | 10.90 |
|  |  |  | Combined | 0-4 | 0.07 |
|  |  |  | Combined | 5-9 | 0.62 |
|  |  |  | Combined | 10-14 | 4.93 |
|  |  |  | Combined | 15-19 | 36.68 |
|  |  |  | Male | 0-19 | 6.95 |
|  |  |  | Female | 0-19 | 15.06 |
|  |  | 2011 | Combined | 0-19 | 10.40 |
|  |  |  | Combined | 0-4 | 0.07 |
|  |  |  | Combined | 5-9 | 0.58 |
|  |  |  | Combined | 10-14 | 4.64 |
|  |  |  | Combined | 15-19 | 34.55 |
|  |  |  | Male | 0-19 | 6.75 |
|  |  |  | Female | 0-19 | 14.24 |
|  |  | 2012 | Combined | 0-19 | 0.98 |
|  |  |  | Combined | 0-4 | 0.09 |
|  |  |  | Combined | 5-9 | 0.48 |
|  |  |  | Combined | 10-14 | 4.62 |
|  |  |  | Combined | 15-19 | 31.94 |
|  |  |  | Male | 0-19 | 6.23 |
|  |  |  | Female | 0-19 | 13.51 |
| Sihvo 2008^a^ [] | Finland | 2000 | Combined | 30 and older | 73.96 |
| Sihvo 2010^a^ [] | Finland | 1994 | Combined | 18 and older | 36.0 |
|  |  | 2003 | Combined | 18 and older | 73.0 |
| Foulon 2010^a^ [] | Finland | 1998 | Combined | 0-19 | 2.23 |
|  |  | 2002 | Combined | 0-19 | 5.24 |
|  |  | 2005 | Combined | 0-19 | 5.93 |
| Autti-Rämö 2011^a^ [] | Finland | 1997 | Combined | 0-26 | 6.4 |
|  |  |  | Combined | 0-6 | 0 |
|  |  |  | Combined | 7-10 | 0.4 |
|  |  |  | Combined | 11-15 | 1.5 |
|  |  |  | Combined | 16-20 | 8.2 |
|  |  |  | Combined | 21-26 | 21 |
|  |  |  | Male | 0-26 | 5.3 |
|  |  |  | Male | 0-6 | 0.1 |
|  |  |  | Male | 7-10 | 0.5 |
|  |  |  | Male | 11-15 | 1.5 |
|  |  |  | Male | 16-20 | 5.5 |
|  |  |  | Male | 21-26 | 17.9 |
|  |  |  | Female | 0-26 | 7.6 |
|  |  |  | Female | 0-6 | 0 |
|  |  |  | Female | 7-10 | 0.4 |
|  |  |  | Female | 11-15 | 1.4 |
|  |  |  | Female | 16-20 | 11 |
|  |  |  | Female | 21-26 | 24.2 |
|  |  | 2002 | Combined | 0-26 | 14 |
|  |  |  | Combined | 0-6 | 0.1 |
|  |  |  | Combined | 7-10 | 0.6 |
|  |  |  | Combined | 11-15 | 3.9 |
|  |  |  | Combined | 16-20 | 21.8 |
|  |  |  | Combined | 21-26 | 38.9 |
|  |  |  | Male | 0-26 | 10.7 |
|  |  |  | Male | 0-6 | 0.1 |
|  |  |  | Male | 7-10 | 0.9 |
|  |  |  | Male | 11-15 | 3.5 |
|  |  |  | Male | 16-20 | 14.1 |
|  |  |  | Male | 21-26 | 31.2 |
|  |  |  | Female | 0-26 | 17.4 |
|  |  |  | Female | 0-6 | 0 |
|  |  |  | Female | 7-10 | 0.3 |
|  |  |  | Female | 11-15 | 4.4 |
|  |  |  | Female | 16-20 | 29.8 |
|  |  |  | Female | 21-26 | 47 |
|  |  | 2007 | Combined | 0-26 | 22.5 |
|  |  |  | Combined | 0-6 | 0 |
|  |  |  | Combined | 7-10 | 0.7 |
|  |  |  | Combined | 11-15 | 5.3 |
|  |  |  | Combined | 16-20 | 31.4 |
|  |  |  | Combined | 21-26 | 64.5 |
|  |  |  | Male | 0-26 | 16 |
|  |  |  | Male | 0-6 | 0 |
|  |  |  | Male | 7-10 | 1.1 |
|  |  |  | Male | 11-15 | 4.6 |
|  |  |  | Male | 16-20 | 19 |
|  |  |  | Male | 21-26 | 47.6 |
|  |  |  | Female | 0-26 | 29.2 |
|  |  |  | Female | 0-6 | 0 |
|  |  |  | Female | 7-10 | 0.4 |
|  |  |  | Female | 11-15 | 5.9 |
|  |  |  | Female | 16-20 | 44.3 |
|  |  |  | Female | 21-26 | 82.2 |
| Zoëga 2009^a^ [] | Iceland | 2003 | Combined | 0-17 | 28.28 |
|  |  | 2004 | Combined | 0-17 | 28.01 |
|  |  | 2005 | Combined | 0-17 | 25.64 |
|  |  | 2006 | Combined | 0-17 | 24.61 |
|  |  | 2007 | Combined | 0-17 | 23.41 |
| Kjosavik 2009^a^ [] | Norway | 2005 | Combined | All | 60 |
| Hartz 2016a^a^ [] | Norway | 2004 | Combined | 13-17 | 6.4 |
|  |  | 2013 | Combined | 13-17 | 9.1 |
|  |  |  | Male | 13-17 | 5.9 |
|  |  |  | Female | 13-17 | 12.4 |
| Hartz 2016b^a^ [] | Norway | 2004 | Male | 0-17 | 2.1 |
|  |  |  | Female | 0-17 | 3.1 |
|  |  | 2005 | Male | 0-17 | 1.7 |
|  |  |  | Female | 0-17 | 2.5 |
|  |  | 2006 | Male | 0-17 | 1.7 |
|  |  |  | Female | 0-17 | 2.4 |
|  |  | 2007 | Male | 0-17 | 1.8 |
|  |  |  | Female | 0-17 | 2.6 |
|  |  | 2008 | Male | 0-17 | 1.8 |
|  |  |  | Female | 0-17 | 2.6 |
|  |  | 2009 | Male | 0-17 | 2 |
|  |  |  | Female | 0-17 | 2.6 |
|  |  | 2010 | Male | 0-17 | 2.2 |
|  |  |  | Female | 0-17 | 2.8 |
|  |  | 2011 | Male | 0-17 | 2.2 |
|  |  |  | Female | 0-17 | 3 |
|  |  | 2012 | Male | 0-17 | 2.2 |
|  |  |  | Female | 0-17 | 3.6 |
|  |  | 2013 | Male | 0-17 | 2.1 |
|  |  |  | Female | 0-17 | 4 |
|  |  | 2014 | Male | 0-17 | 2 |
|  |  |  | Female | 0-17 | 10 |
| Gómez-Lumbreras 2021^a^ [] | Norway | 2008 | Combined | 0-19 | 4.17 |
|  |  |  | Male | 0-19 | 2.95 |
|  |  |  | Female | 0-19 | 5.46 |
|  |  | 2009 | Combined | 0-19 | 4.26 |
|  |  |  | Male | 0-19 | 3.12 |
|  |  |  | Female | 0-19 | 5.47 |
|  |  | 2010 | Combined | 0-19 | 4.35 |
|  |  |  | Male | 0-19 | 3.14 |
|  |  |  | Female | 0-19 | 5.61 |
|  |  | 2011 | Combined | 0-19 | 4.68 |
|  |  |  | Male | 0-19 | 3.23 |
|  |  |  | Female | 0-19 | 6.19 |
|  |  | 2012 | Combined | 0-19 | 5.17 |
|  |  |  | Male | 0-19 | 3.5 |
|  |  |  | Female | 0-19 | 6.92 |
|  |  | 2013 | Combined | 0-19 | 5.35 |
|  |  |  | Male | 0-19 | 3.5 |
|  |  |  | Female | 0-19 | 7.3 |
|  |  | 2014 | Combined | 0-19 | 5.52 |
|  |  |  | Male | 0-19 | 3.45 |
|  |  |  | Female | 0-19 | 7.7 |
|  |  | 2015 | Combined | 0-19 | 5.8 |
|  |  |  | Male | 0-19 | 3.57 |
|  |  |  | Female | 0-19 | 8.14 |
|  |  | 2016 | Combined | 0-19 | 5.86 |
|  |  |  | Male | 0-19 | 3.5 |
|  |  |  | Female | 0-19 | 8.35 |
|  |  | 2017 | Combined | 0-19 | 5.95 |
|  |  |  | Male | 0-19 | 3.58 |
|  |  |  | Male | 0-4 | 0 |
|  |  |  | Male | 5-9 | 0.24 |
|  |  |  | Male | 10-14 | 1.74 |
|  |  |  | Male | 15-19 | 11.68 |
|  |  |  | Female | 0-19 | 8.45 |
|  |  |  | Female | 0-4 | 0 |
|  |  |  | Female | 5-9 | 0.12 |
|  |  |  | Female | 10-14 | 1.8 |
|  |  |  | Female | 15-19 | 30.54 |
| Lien 2023^a^ [] | Norway | 2004 | Combined | 15-19 | 9.92 |
|  |  |  | Male | 15-19 | 7.23 |
|  |  |  | Female | 15-19 | 12.69 |
|  |  | 2005 | Combined | 15-19 | 9.29 |
|  |  |  | Male | 15-19 | 6.28 |
|  |  |  | Female | 15-19 | 12.34 |
|  |  | 2006 | Combined | 15-19 | 9.86 |
|  |  |  | Male | 15-19 | 7.18 |
|  |  |  | Female | 15-19 | 12.56 |
|  |  | 2007 | Combined | 15-19 | 10.42 |
|  |  |  | Male | 15-19 | 7.15 |
|  |  |  | Female | 15-19 | 13.73 |
|  |  | 2008 | Combined | 15-19 | 10.07 |
|  |  |  | Male | 15-19 | 7.04 |
|  |  |  | Female | 15-19 | 13.13 |
|  |  | 2009 | Combined | 15-19 | 10.20 |
|  |  |  | Male | 15-19 | 7.36 |
|  |  |  | Female | 15-19 | 13.13 |
|  |  | 2010 | Combined | 15-19 | 10.53 |
|  |  |  | Male | 15-19 | 7.43 |
|  |  |  | Female | 15-19 | 13.79 |
|  |  | 2011 | Combined | 15-19 | 11.24 |
|  |  |  | Male | 15-19 | 7.96 |
|  |  |  | Female | 15-19 | 14.69 |
|  |  | 2012 | Combined | 15-19 | 12.27 |
|  |  |  | Male | 15-19 | 7.07 |
|  |  |  | Female | 15-19 | 17.72 |
|  |  | 2013 | Combined | 15-19 | 14.45 |
|  |  |  | Male | 15-19 | 9.17 |
|  |  |  | Female | 15-19 | 20.00 |
|  |  | 2014 | Combined | 15-19 | 14.06 |
|  |  |  | Male | 15-19 | 9.30 |
|  |  |  | Female | 15-19 | 19.06 |
|  |  | 2015 | Combined | 15-19 | 14.83 |
|  |  |  | Male | 15-19 | 8.90 |
|  |  |  | Female | 15-19 | 20.96 |
|  |  | 2016 | Combined | 15-19 | 15.90 |
|  |  |  | Male | 15-19 | 9.80 |
|  |  |  | Female | 15-19 | 22.18 |
|  |  | 2017 | Combined | 15-19 | 16.18 |
|  |  |  | Male | 15-19 | 9.59 |
|  |  |  | Female | 15-19 | 22.98 |
|  |  | 2018 | Combined | 15-19 | 16.95 |
|  |  |  | Male | 15-19 | 10.70 |
|  |  |  | Female | 15-19 | 23.42 |
|  |  | 2019 | Combined | 15-19 | 16.28 |
|  |  |  | Male | 15-19 | 9.11 |
|  |  |  | Female | 15-19 | 23.58 |
|  |  | 2020 | Combined | 15-19 | 16.97 |
|  |  |  | Male | 15-19 | 9.97 |
|  |  |  | Female | 15-19 | 24.07 |
| Steffenak 2012^a^ [] | Norway | 2006 | Male | 15-16 | 4.30 |
|  |  |  | Female | 15-16 | 7.80 |
|  |  | 2008 | Male | 15-16 | 5.00 |
|  |  |  | Female | 15-16 | 8.20 |
|  |  | 2010 | Male | 15-16 | 5.20 |
|  |  |  | Female | 15-16 | 9.00 |
| Wesselhoeft 2020^a^ [] | Norway | 2007 | Combined | 5-19 | 5.08 |
|  |  | 2008 | Combined | 5-19 | 5.37 |
|  |  | 2009 | Combined | 5-19 | 5.5 |
|  |  | 2010 | Combined | 5-19 | 5.61 |
|  |  | 2011 | Combined | 5-19 | 6.05 |
|  |  | 2012 | Combined | 5-19 | 6.71 |
|  |  | 2013 | Combined | 5-19 | 6.96 |
|  |  | 2014 | Combined | 5-19 | 7.15 |
|  |  | 2015 | Combined | 5-19 | 7.48 |
|  |  | 2016 | Combined | 5-19 | 7.53 |
|  |  | 2017 | Combined | 5-19 | 7.6 |
|  |  | 2007 | Female | 5-9 | 0.09 |
|  |  | 2007 | Male | 5-9 | 0.28 |
|  |  | 2007 | Female | 10-14 | 1.06 |
|  |  | 2007 | Male | 10-14 | 1.59 |
|  |  | 2007 | Female | 15-19 | 18.62 |
|  |  | 2007 | Male | 15-19 | 8.98 |
|  |  | 2008 | Female | 5-9 | 0.09 |
|  |  | 2008 | Male | 5-9 | 0.32 |
|  |  | 2008 | Female | 10-14 | 1.24 |
|  |  | 2008 | Male | 10-14 | 1.74 |
|  |  | 2008 | Female | 15-19 | 19.42 |
|  |  | 2008 | Male | 15-19 | 9.10 |
|  |  | 2009 | Female | 5-9 | 0.10 |
|  |  | 2009 | Male | 5-9 | 0.24 |
|  |  | 2009 | Female | 10-14 | 1.36 |
|  |  | 2009 | Male | 10-14 | 1.89 |
|  |  | 2009 | Female | 15-19 | 19.21 |
|  |  | 2009 | Male | 15-19 | 9.67 |
|  |  | 2010 | Female | 5-9 | 0.16 |
|  |  | 2010 | Male | 5-9 | 0.30 |
|  |  | 2010 | Female | 10-14 | 1.58 |
|  |  | 2010 | Male | 10-14 | 1.91 |
|  |  | 2010 | Female | 15-19 | 19.43 |
|  |  | 2010 | Male | 15-19 | 9.63 |
|  |  | 2011 | Female | 5-9 | 0.14 |
|  |  | 2011 | Male | 5-9 | 0.24 |
|  |  | 2011 | Female | 10-14 | 1.47 |
|  |  | 2011 | Male | 10-14 | 2.08 |
|  |  | 2011 | Female | 15-19 | 21.79 |
|  |  | 2011 | Male | 15-19 | 9.82 |
|  |  | 2012 | Female | 5-9 | 0.10 |
|  |  | 2012 | Male | 5-9 | 0.18 |
|  |  | 2012 | Female | 10-14 | 1.74 |
|  |  | 2012 | Male | 10-14 | 2.29 |
|  |  | 2012 | Female | 15-19 | 24.36 |
|  |  | 2012 | Male | 15-19 | 10.70 |
|  |  | 2013 | Female | 5-9 | 0.09 |
|  |  | 2013 | Male | 5-9 | 0.23 |
|  |  | 2013 | Female | 10-14 | 1.79 |
|  |  | 2013 | Male | 10-14 | 2.03 |
|  |  | 2013 | Female | 15-19 | 25.72 |
|  |  | 2013 | Male | 15-19 | 10.94 |
|  |  | 2014 | Female | 5-9 | 0.10 |
|  |  | 2014 | Male | 5-9 | 0.21 |
|  |  | 2014 | Female | 10-14 | 1.80 |
|  |  | 2014 | Male | 10-14 | 1.92 |
|  |  | 2014 | Female | 15-19 | 27.26 |
|  |  | 2014 | Male | 15-19 | 10.90 |
|  |  | 2015 | Female | 5-9 | 0.08 |
|  |  | 2015 | Male | 5-9 | 0.27 |
|  |  | 2015 | Female | 10-14 | 1.92 |
|  |  | 2015 | Male | 10-14 | 1.97 |
|  |  | 2015 | Female | 15-19 | 28.84 |
|  |  | 2015 | Male | 15-19 | 11.25 |
|  |  | 2016 | Female | 5-9 | 0.08 |
|  |  | 2016 | Male | 5-9 | 0.19 |
|  |  | 2016 | Female | 10-14 | 1.81 |
|  |  | 2016 | Male | 10-14 | 1.88 |
|  |  | 2016 | Female | 15-19 | 29.72 |
|  |  | 2016 | Male | 15-19 | 11.14 |
|  |  | 2017 | Female | 5-9 | 0.12 |
|  |  | 2017 | Male | 5-9 | 0.24 |
|  |  | 2017 | Female | 10-14 | 1.82 |
|  |  | 2017 | Male | 10-14 | 1.78 |
|  |  | 2017 | Female | 15-19 | 30.19 |
|  |  | 2017 | Male | 15-19 | 11.49 |
| Bojanić 2024^a^ [] | Norway | 2006 | Combined | 5 and older | 60.00 |
|  |  |  | Combined | 5-19 | 4.60 |
|  |  |  | Combined | 20-64 | 68.20 |
|  |  |  | Combined | 65 and older | 107.40 |
|  |  |  | Male | 15-19 | 7.80 |
|  |  |  | Male | 20-44 | 40.90 |
|  |  |  | Male | 45-64 | 63.40 |
|  |  |  | Male | 65-74 | 65.20 |
|  |  |  | Male | 75 and older | 89.00 |
|  |  |  | Female | 15-19 | 17.70 |
|  |  |  | Female | 20-44 | 69.20 |
|  |  |  | Female | 45-64 | 120.60 |
|  |  |  | Female | 65-74 | 132.00 |
|  |  |  | Female | 75 and older | 140.60 |
|  |  | 2009 | Combined | 5 and older | 60.60 |
|  |  |  | Combined | 5-19 | 5.50 |
|  |  |  | Combined | 20-64 | 70.80 |
|  |  |  | Combined | 65 and older | 112.30 |
|  |  |  | Male | 15-19 | 9.60 |
|  |  |  | Male | 20-44 | 41.30 |
|  |  |  | Male | 45-64 | 63.60 |
|  |  |  | Male | 65-74 | 64.40 |
|  |  |  | Male | 75 and older | 90.80 |
|  |  |  | Female | 15-19 | 19.50 |
|  |  |  | Female | 20-44 | 67.80 |
|  |  |  | Female | 45-64 | 120.60 |
|  |  |  | Female | 65-74 | 131.44 |
|  |  |  | Female | 75 and older | 147.10 |
|  |  | 2012 | Combined | 5 and older | 61.80 |
|  |  |  | Combined | 5-19 | 6.70 |
|  |  |  | Combined | 20-64 | 71.10 |
|  |  |  | Combined | 65 and older | 112.50 |
|  |  |  | Male | 15-19 | 10.70 |
|  |  |  | Male | 20-44 | 41.00 |
|  |  |  | Male | 45-64 | 63.70 |
|  |  |  | Male | 65-74 | 65.70 |
|  |  |  | Male | 75 and older | 91.30 |
|  |  |  | Female | 15-19 | 24.30 |
|  |  |  | Female | 20-44 | 70.70 |
|  |  |  | Female | 45-64 | 121.00 |
|  |  |  | Female | 65-74 | 132.20 |
|  |  |  | Female | 75 and older | 153.20 |
|  |  | 2015 | Combined | 5 and older | 62.20 |
|  |  |  | Combined | 5-19 | 7.50 |
|  |  |  | Combined | 20-64 | 70.90 |
|  |  |  | Combined | 65 and older | 110.30 |
|  |  |  | Male | 15-19 | 11.30 |
|  |  |  | Male | 20-44 | 40.60 |
|  |  |  | Male | 45-64 | 63.00 |
|  |  |  | Male | 65-74 | 66.40 |
|  |  |  | Male | 75 and older | 87.50 |
|  |  |  | Female | 15-19 | 28.90 |
|  |  |  | Female | 20-44 | 71.60 |
|  |  |  | Female | 45-64 | 121.10 |
|  |  |  | Female | 65-74 | 130.70 |
|  |  |  | Female | 75 and older | 151.50 |
|  |  | 2018 | Combined | 5 and older | 62.70 |
|  |  |  | Combined | 5-19 | 7.30 |
|  |  |  | Combined | 20-64 | 71.50 |
|  |  |  | Combined | 65 and older | 107.50 |
|  |  |  | Male | 15-19 | 12.40 |
|  |  |  | Male | 20-44 | 41.10 |
|  |  |  | Male | 45-64 | 62.00 |
|  |  |  | Male | 65-74 | 66.30 |
|  |  |  | Male | 75 and older | 84.00 |
|  |  |  | Female | 15-19 | 28.40 |
|  |  |  | Female | 20-44 | 74.80 |
|  |  |  | Female | 45-64 | 118.80 |
|  |  |  | Female | 65-74 | 130.40 |
|  |  |  | Female | 75 and older | 147.60 |
|  |  | 2021 | Combined | 5 and older | 68.90 |
|  |  |  | Combined | 5-19 | 8.20 |
|  |  |  | Combined | 20-64 | 75.10 |
|  |  |  | Combined | 65 and older | 107.70 |
|  |  |  | Male | 15-19 | 12.50 |
|  |  |  | Male | 20-44 | 44.00 |
|  |  |  | Male | 45-64 | 63.10 |
|  |  |  | Male | 65-74 | 67.90 |
|  |  |  | Male | 75 and older | 83.80 |
|  |  |  | Female | 15-19 | 31.60 |
|  |  |  | Female | 20-44 | 81.50 |
|  |  |  | Female | 45-64 | 121.70 |
|  |  |  | Female | 65-74 | 131.60 |
|  |  |  | Female | 75 and older | 146.80 |
| Gómez-Lumbreras 2021^a^ [] | Sweden | 2008 | Combined | 0-19 | 7.18 |
|  |  |  | Male | 0-19 | 5.06 |
|  |  |  | Female | 0-19 | 9.43 |
|  |  | 2009 | Combined | 0-19 | 7.45 |
|  |  |  | Male | 0-19 | 5.37 |
|  |  |  | Female | 0-19 | 9.66 |
|  |  | 2010 | Combined | 0-19 | 7.92 |
|  |  |  | Male | 0-19 | 5.8 |
|  |  |  | Female | 0-19 | 10.17 |
|  |  | 2011 | Combined | 0-19 | 8.47 |
|  |  |  | Male | 0-19 | 6.29 |
|  |  |  | Female | 0-19 | 10.79 |
|  |  | 2012 | Combined | 0-19 | 9.1 |
|  |  |  | Male | 0-19 | 6.77 |
|  |  |  | Female | 0-19 | 11.58 |
|  |  | 2013 | Combined | 0-19 | 9.77 |
|  |  |  | Male | 0-19 | 7.17 |
|  |  |  | Female | 0-19 | 12.53 |
|  |  | 2014 | Combined | 0-19 | 10.41 |
|  |  |  | Male | 0-19 | 7.62 |
|  |  |  | Female | 0-19 | 13.37 |
|  |  | 2015 | Combined | 0-19 | 11.41 |
|  |  |  | Male | 0-19 | 8.19 |
|  |  |  | Female | 0-19 | 14.83 |
|  |  | 2016 | Combined | 0-19 | 12.56 |
|  |  |  | Male | 0-19 | 8.6 |
|  |  |  | Female | 0-19 | 16.77 |
|  |  | 2017 | Combined | 0-19 | 13.68 |
|  |  |  | Male | 0-19 | 9.33 |
|  |  |  | Male | 0-4 | 0.023 |
|  |  |  | Male | 5-9 | 0.86 |
|  |  |  | Male | 10-14 | 9 |
|  |  |  | Male | 15-19 | 29.35 |
|  |  |  | Female | 0-19 | 18.33 |
|  |  |  | Female | 0-4 | 0.42 |
|  |  |  | Female | 5-9 | 0.42 |
|  |  |  | Female | 10-14 | 9.66 |
|  |  |  | Female | 15-19 | 69.35 |
| Wesselhoeft 2020 ^a^ [] | Sweden | 2007 | Combined | 5-19 | 8.98 |
|  |  | 2008 | Combined | 5-19 | 9.5 |
|  |  | 2009 | Combined | 5-19 | 9.93 |
|  |  | 2010 | Combined | 5-19 | 10.68 |
|  |  | 2011 | Combined | 5-19 | 11.49 |
|  |  | 2012 | Combined | 5-19 | 12.36 |
|  |  | 2013 | Combined | 5-19 | 13.2 |
|  |  | 2014 | Combined | 5-19 | 14.03 |
|  |  | 2015 | Combined | 5-19 | 15.24 |
|  |  | 2016 | Combined | 5-19 | 16.61 |
|  |  | 2017 | Combined | 5-19 | 18.03 |
|  |  | 2007 | Female | 5-9 | 0.17 |
|  |  | 2007 | Male | 5-9 | 0.31 |
|  |  | 2007 | Female | 10-14 | 3.03 |
|  |  | 2007 | Male | 10-14 | 3.39 |
|  |  | 2007 | Female | 15-19 | 28.17 |
|  |  | 2007 | Male | 15-19 | 13.04 |
|  |  | 2008 | Female | 5-9 | 0.19 |
|  |  | 2008 | Male | 5-9 | 0.32 |
|  |  | 2008 | Female | 10-14 | 3.27 |
|  |  | 2008 | Male | 10-14 | 3.71 |
|  |  | 2008 | Female | 15-19 | 29.29 |
|  |  | 2008 | Male | 15-19 | 13.91 |
|  |  | 2009 | Female | 5-9 | 0.15 |
|  |  | 2009 | Male | 5-9 | 0.36 |
|  |  | 2009 | Female | 10-14 | 3.35 |
|  |  | 2009 | Male | 10-14 | 3.63 |
|  |  | 2009 | Female | 15-19 | 30.33 |
|  |  | 2009 | Male | 15-19 | 15.23 |
|  |  | 2010 | Female | 5-9 | 0.22 |
|  |  | 2010 | Male | 5-9 | 0.47 |
|  |  | 2010 | Female | 10-14 | 3.71 |
|  |  | 2010 | Male | 10-14 | 4.21 |
|  |  | 2010 | Female | 15-19 | 32.90 |
|  |  | 2010 | Male | 15-19 | 16.75 |
|  |  | 2011 | Female | 5-9 | 0.26 |
|  |  | 2011 | Male | 5-9 | 0.52 |
|  |  | 2011 | Female | 10-14 | 4.29 |
|  |  | 2011 | Male | 10-14 | 4.81 |
|  |  | 2011 | Female | 15-19 | 36.04 |
|  |  | 2011 | Male | 15-19 | 18.72 |
|  |  | 2012 | Female | 5-9 | 0.28 |
|  |  | 2012 | Male | 5-9 | 0.66 |
|  |  | 2012 | Female | 10-14 | 4.95 |
|  |  | 2012 | Male | 10-14 | 5.56 |
|  |  | 2012 | Female | 15-19 | 40.12 |
|  |  | 2012 | Male | 15-19 | 20.53 |
|  |  | 2013 | Female | 5-9 | 0.29 |
|  |  | 2013 | Male | 5-9 | 0.63 |
|  |  | 2013 | Female | 10-14 | 5.46 |
|  |  | 2013 | Male | 10-14 | 6.20 |
|  |  | 2013 | Female | 15-19 | 44.96 |
|  |  | 2013 | Male | 15-19 | 22.14 |
|  |  | 2014 | Female | 5-9 | 0.36 |
|  |  | 2014 | Male | 5-9 | 0.67 |
|  |  | 2014 | Female | 10-14 | 6.33 |
|  |  | 2014 | Male | 10-14 | 6.79 |
|  |  | 2014 | Female | 15-19 | 49.15 |
|  |  | 2014 | Male | 15-19 | 23.99 |
|  |  | 2015 | Female | 5-9 | 0.40 |
|  |  | 2015 | Male | 5-9 | 0.77 |
|  |  | 2015 | Female | 10-14 | 7.33 |
|  |  | 2015 | Male | 10-14 | 7.57 |
|  |  | 2015 | Female | 15-19 | 55.05 |
|  |  | 2015 | Male | 15-19 | 25.73 |
|  |  | 2016 | Female | 5-9 | 0.41 |
|  |  | 2016 | Male | 5-9 | 0.80 |
|  |  | 2016 | Female | 10-14 | 8.70 |
|  |  | 2016 | Male | 10-14 | 8.34 |
|  |  | 2016 | Female | 15-19 | 62.05 |
|  |  | 2016 | Male | 15-19 | 26.28 |
|  |  | 2017 | Female | 5-9 | 0.42 |
|  |  | 2017 | Male | 5-9 | 0.94 |
|  |  | 2017 | Female | 10-14 | 9.38 |
|  |  | 2017 | Male | 10-14 | 8.72 |
|  |  | 2017 | Female | 15-19 | 67.81 |
|  |  | 2017 | Male | 15-19 | 28.38 |
| Bojanić 2024 ^a^ [] | Sweden | 2006 | Combined | 5 and older | 79.7 |
|  |  |  | Combined | 5-19 | 8 |
|  |  |  | Combined | 20-64 | 88.1 |
|  |  |  | Combined | 65 and older | 152.6 |
|  |  |  | Male | 15-19 | 12.1 |
|  |  |  | Male | 20-44 | 48.4 |
|  |  |  | Male | 45-64 | 74.2 |
|  |  |  | Male | 65-74 | 77.9 |
|  |  |  | Male | 75 and older | 151.5 |
|  |  |  | Female | 15-19 | 26.8 |
|  |  |  | Female | 20-44 | 91.8 |
|  |  |  | Female | 45-64 | 145.2 |
|  |  |  | Female | 65-74 | 139.5 |
|  |  |  | Female | 75 and older | 229.7 |
|  |  | 2009 | Combined | 5 and older | 79.9 |
|  |  |  | Combined | 5-19 | 9.9 |
|  |  |  | Combined | 20-64 | 86.8 |
|  |  |  | Combined | 65 and older | 153.4 |
|  |  |  | Male | 15-19 | 15.2 |
|  |  |  | Male | 20-44 | 49.1 |
|  |  |  | Male | 45-64 | 74 |
|  |  |  | Male | 65-74 | 80.6 |
|  |  |  | Male | 75 and older | 150.3 |
|  |  |  | Female | 15-19 | 30.2 |
|  |  |  | Female | 20-44 | 91.1 |
|  |  |  | Female | 45-64 | 141.1 |
|  |  |  | Female | 65-74 | 144.8 |
|  |  |  | Female | 75 and older | 231.8 |
|  |  | 2012 | Combined | 5 and older | 85.3 |
|  |  |  | Combined | 5-19 | 12.3 |
|  |  |  | Combined | 20-64 | 92.3 |
|  |  |  | Combined | 65 and older | 156.4 |
|  |  |  | Male | 15-19 | 19.6 |
|  |  |  | Male | 20-44 | 54.1 |
|  |  |  | Male | 45-64 | 78.1 |
|  |  |  | Male | 65-74 | 84.2 |
|  |  |  | Male | 75 and older | 155.6 |
|  |  |  | Female | 15-19 | 38.2 |
|  |  |  | Female | 20-44 | 100.5 |
|  |  |  | Female | 45-64 | 147 |
|  |  |  | Female | 65-74 | 152.5 |
|  |  |  | Female | 75 and older | 240.2 |
|  |  | 2015 | Combined | 5 and older | 94.2 |
|  |  |  | Combined | 5-19 | 15.5 |
|  |  |  | Combined | 20-64 | 104.2 |
|  |  |  | Combined | 65 and older | 160.8 |
|  |  |  | Male | 15-19 | 25.5 |
|  |  |  | Male | 20-44 | 63.6 |
|  |  |  | Male | 45-64 | 85.2 |
|  |  |  | Male | 65-74 | 88.6 |
|  |  |  | Male | 75 and older | 162.9 |
|  |  |  | Female | 15-19 | 54.5 |
|  |  |  | Female | 20-44 | 119 |
|  |  |  | Female | 45-64 | 160.5 |
|  |  |  | Female | 65-74 | 157.8 |
|  |  |  | Female | 75 and older | 250.3 |
|  |  | 2018 | Combined | 5 and older | 99.2 |
|  |  |  | Combined | 5-19 | 19.7 |
|  |  |  | Combined | 20-64 | 110.4 |
|  |  |  | Combined | 65 and older | 165.8 |
|  |  |  | Male | 15-19 | 31.7 |
|  |  |  | Male | 20-44 | 67.9 |
|  |  |  | Male | 45-64 | 88.3 |
|  |  |  | Male | 65-74 | 92.4 |
|  |  |  | Male | 75 and older | 169.7 |
|  |  |  | Female | 15-19 | 72.7 |
|  |  |  | Female | 20-44 | 130.7 |
|  |  |  | Female | 45-64 | 165.6 |
|  |  |  | Female | 65-74 | 159.6 |
|  |  |  | Female | 75 and older | 259.4 |
|  |  | 2021 | Combined | 5 and older | 106.9 |
|  |  |  | Combined | 5-19 | 24.3 |
|  |  |  | Combined | 20-64 | 119.2 |
|  |  |  | Combined | 65 and older | 174.8 |
|  |  |  | Male | 15-19 | 34.7 |
|  |  |  | Male | 20-44 | 74.1 |
|  |  |  | Male | 45-64 | 93.2 |
|  |  |  | Male | 65-74 | 98.4 |
|  |  |  | Male | 75 and older | 176.1 |
|  |  |  | Female | 15-19 | 87.9 |
|  |  |  | Female | 20-44 | 144.1 |
|  |  |  | Female | 45-64 | 175.1 |
|  |  |  | Female | 65-74 | 166.5 |
|  |  |  | Female | 75 and older | 272.4 |
| Lagerberg 2019 ^a^ [] | Sweden | 2006 | Combined | 0-24 | 14 |
|  |  |  | Male | 0-24 | 9 |
|  |  |  | Male | 0-11 | 0.5 |
|  |  |  | Male | 12-17 | 6 |
|  |  |  | Male | 18-24 | 25 |
|  |  |  | Female | 0-24 | 18 |
|  |  |  | Female | 0-11 | 0.2 |
|  |  |  | Female | 12-17 | 11 |
|  |  |  | Female | 18-24 | 52 |
|  |  | 2007 | Combined | 0-24 | 14 |
|  |  |  | Male | 0-24 | 10 |
|  |  |  | Male | 0-11 | 0.4 |
|  |  |  | Male | 12-17 | 7 |
|  |  |  | Male | 18-24 | 26 |
|  |  |  | Female | 0-24 | 19 |
|  |  |  | Female | 0-11 | 0.2 |
|  |  |  | Female | 12-17 | 13 |
|  |  |  | Female | 18-24 | 54 |
|  |  | 2008 | Combined | 0-24 | 15 |
|  |  |  | Male | 0-24 | 10 |
|  |  |  | Male | 0-11 | 0.4 |
|  |  |  | Male | 12-17 | 8 |
|  |  |  | Male | 18-24 | 27 |
|  |  |  | Female | 0-24 | 20 |
|  |  |  | Female | 0-11 | 0.2 |
|  |  |  | Female | 12-17 | 13 |
|  |  |  | Female | 18-24 | 54 |
|  |  | 2009 | Combined | 0-24 | 16 |
|  |  |  | Male | 0-24 | 11 |
|  |  |  | Male | 0-11 | 0.5 |
|  |  |  | Male | 12-17 | 8 |
|  |  |  | Male | 18-24 | 28 |
|  |  |  | Female | 0-24 | 21 |
|  |  |  | Female | 0-11 | 0.2 |
|  |  |  | Female | 12-17 | 14 |
|  |  |  | Female | 18-24 | 55 |
|  |  | 2010 | Combined | 0-24 | 17 |
|  |  |  | Male | 0-24 | 12 |
|  |  |  | Male | 0-11 | 0.6 |
|  |  |  | Male | 12-17 | 9 |
|  |  |  | Male | 18-24 | 29 |
|  |  |  | Female | 0-24 | 22 |
|  |  |  | Female | 0-11 | 0.3 |
|  |  |  | Female | 12-17 | 15 |
|  |  |  | Female | 18-24 | 57 |
|  |  | 2011 | Combined | 0-24 | 18 |
|  |  |  | Male | 0-24 | 13 |
|  |  |  | Male | 0-11 | 0.6 |
|  |  |  | Male | 12-17 | 11 |
|  |  |  | Male | 18-24 | 32 |
|  |  |  | Female | 0-24 | 23 |
|  |  |  | Female | 0-11 | 0.3 |
|  |  |  | Female | 12-17 | 17 |
|  |  |  | Female | 18-24 | 60 |
|  |  | 2012 | Combined | 0-24 | 19 |
|  |  |  | Male | 0-24 | 14 |
|  |  |  | Male | 0-11 | 0.8 |
|  |  |  | Male | 12-17 | 11 |
|  |  |  | Male | 18-24 | 34 |
|  |  |  | Female | 0-24 | 25 |
|  |  |  | Female | 0-11 | 0.4 |
|  |  |  | Female | 12-17 | 19 |
|  |  |  | Female | 18-24 | 65 |
|  |  | 2013 | Combined | 0-24 | 21 |
|  |  |  | Male | 0-24 | 15 |
|  |  |  | Male | 0-11 | 0.8 |
|  |  |  | Male | 12-17 | 13 |
|  |  |  | Male | 18-24 | 36 |
|  |  |  | Female | 0-24 | 27 |
|  |  |  | Female | 0-11 | 0.4 |
|  |  |  | Female | 12-17 | 21 |
|  |  |  | Female | 18-24 | 70 |
| Forslund 2020 ^a^ [] | Sweden | 2011 | Combined | 20 and older | 82.36 |
|  |  | 2011 | Female | 20-29 | 63.45 |
|  |  | 2011 | Female | 30-44 | 93.14 |
|  |  | 2011 | Female | 45-64 | 120.83 |
|  |  | 2011 | Female | 65 and older | 140.39 |
|  |  | 2011 | Male | 20-29 | 34.30 |
|  |  | 2011 | Male | 30-44 | 50.01 |
|  |  | 2011 | Male | 45-64 | 64.82 |
|  |  | 2011 | Male | 65 and older | 76.01 |
|  |  | 2012 | Combined | 20 and older | 83.96 |
|  |  | 2012 | Female | 20-29 | 67.47 |
|  |  | 2012 | Female | 30-44 | 96.92 |
|  |  | 2012 | Female | 45-64 | 122.40 |
|  |  | 2012 | Female | 65 and older | 141.46 |
|  |  | 2012 | Male | 20-29 | 35.89 |
|  |  | 2012 | Male | 30-44 | 50.65 |
|  |  | 2012 | Male | 45-64 | 65.55 |
|  |  | 2012 | Male | 65 and older | 75.90 |
|  |  | 2013 | Combined | 20 and older | 86.78 |
|  |  | 2013 | Female | 20-29 | 73.03 |
|  |  | 2013 | Female | 30-44 | 101.92 |
|  |  | 2013 | Female | 45-64 | 124.76 |
|  |  | 2013 | Female | 65 and older | 143.38 |
|  |  | 2013 | Male | 20-29 | 38.46 |
|  |  | 2013 | Male | 30-44 | 53.10 |
|  |  | 2013 | Male | 45-64 | 66.85 |
|  |  | 2013 | Male | 65 and older | 78.17 |
|  |  | 2014 | Combined | 20 and older | 89.85 |
|  |  | 2014 | Female | 20-29 | 77.01 |
|  |  | 2014 | Female | 30-44 | 107.33 |
|  |  | 2014 | Female | 45-64 | 129.03 |
|  |  | 2014 | Female | 65 and older | 146.28 |
|  |  | 2014 | Male | 20-29 | 40.91 |
|  |  | 2014 | Male | 30-44 | 54.83 |
|  |  | 2014 | Male | 45-64 | 68.82 |
|  |  | 2014 | Male | 65 and older | 79.94 |
|  |  | 2015 | Combined | 20 and older | 93.41 |
|  |  | 2015 | Female | 20-29 | 82.51 |
|  |  | 2015 | Female | 30-44 | 113.40 |
|  |  | 2015 | Female | 45-64 | 132.15 |
|  |  | 2015 | Female | 65 and older | 149.22 |
|  |  | 2015 | Male | 20-29 | 44.41 |
|  |  | 2015 | Male | 30-44 | 57.64 |
|  |  | 2015 | Male | 45-64 | 71.19 |
|  |  | 2015 | Male | 65 and older | 82.09 |
|  |  | 2016 | Combined | 20 and older | 95.16 |
|  |  | 2016 | Female | 20-29 | 87.60 |
|  |  | 2016 | Female | 30-44 | 116.86 |
|  |  | 2016 | Female | 45-64 | 134.25 |
|  |  | 2016 | Female | 65 and older | 149.28 |
|  |  | 2016 | Male | 20-29 | 46.25 |
|  |  | 2016 | Male | 30-44 | 58.70 |
|  |  | 2016 | Male | 45-64 | 71.45 |
|  |  | 2016 | Male | 65 and older | 82.98 |
|  |  | 2017 | Combined | 20 and older | 95.41 |
|  |  | 2017 | Female | 20-29 | 90.31 |
|  |  | 2017 | Female | 30-44 | 118.82 |
|  |  | 2017 | Female | 45-64 | 133.53 |
|  |  | 2017 | Female | 65 and older | 149.82 |
|  |  | 2017 | Male | 20-29 | 46.33 |
|  |  | 2017 | Male | 30-44 | 58.41 |
|  |  | 2017 | Male | 45-64 | 70.15 |
|  |  | 2017 | Male | 65 and older | 83.57 |
| Loikas 2013 ^a^ [] | Sweden | 2010 | Male | All ages | 55.35 |
|  |  |  | Male | 0-4 | 0.01 |
|  |  |  | Male | 5-14 | 2.29 |
|  |  |  | Male | 15-44 | 44.71 |
|  |  |  | Male | 45-64 | 75.05 |
|  |  |  | Male | 65-74 | 80.18 |
|  |  |  | Male | 75-84 | 123.87 |
|  |  |  | Male | 85 and older | 202.83 |
|  |  |  | Female | All ages | 106.6 |
|  |  |  | Female | 0-4 | 0.02 |
|  |  |  | Female | 5-14 | 1.91 |
|  |  |  | Female | 15-44 | 82.92 |
|  |  |  | Female | 45-64 | 142.07 |
|  |  |  | Female | 65-74 | 146.95 |
|  |  |  | Female | 75-84 | 196.93 |
|  |  |  | Female | 85 and older | 307.37 |
| Wastesson 2012^b^ [] | Sweden | 2008 | Combined | 80-89 | 160.22 |
|  |  |  | Combined | 90-99 | 206.79 |
|  |  |  | Combined | 100 and older | 187.46 |
|  |  |  | Combined | 80 and older | 168.04 |
| Corneliusson 2024 ^b^ [] | Sweden | 2000-2002 | Combined | 85 and older | 180.9 |
|  |  |  | Male | 85 and older | 121.5 |
|  |  |  | Female | 85 and older | 204.5 |
|  |  | 2005-2007 | Combined | 85 and older | 174.8 |
|  |  |  | Male | 85 and older | 113.5 |
|  |  |  | Female | 85 and older | 201.2 |
|  |  | 2010-2012 | Combined | 85 and older | 220.2 |
|  |  |  | Male | 85 and older | 175.9 |
|  |  |  | Female | 85 and older | 242.4 |
|  |  | 2015-2017 | Combined | 85 and older | 247.1 |
|  |  |  | Male | 85 and older | 196.3 |
|  |  |  | Female | 85 and older | 270.7 |
| ^a^Incidence per 1000 person-years.  ^b^Incidence per 1000 population. | | | | | |
