## Supplemental table 4 for "Age-sex specific prevalence and incidence of antidepressant prescribing in the Nordic countries: a systematic review"

| **Supplemental table 4.** Results in studies on incidence of antidepressant prescription. | | | | | | |
| --- | --- | --- | --- | --- | --- | --- |
| **Study** | **Country** | **Year** | **Sex** | **Ages** | **Incidence per 1000** | **Definition of incidence** |
| Pottegård 2014^a^ [] | Denmark | 1997 | Combined | 5-17 | 0.57 | First-ever dispensation |
|  |  | 2010 | Combined | 5-17 | 3.3 |  |
|  |  | 2011 | Combined | 5-17 | 2.55 |  |
| Skovlund 2017^a^ [] | Denmark | 1996 | Male | 10-11 | 0.5 | First-ever dispensation |
|  |  |  | Male | 12-14 | 0.8 |  |
|  |  |  | Male | 15-19 | 1.5 |  |
|  |  |  | Male | 20-24 | 3.7 |  |
|  |  |  | Male | 25-29 | 5.5 |  |
|  |  |  | Male | 30-34 | 7.6 |  |
|  |  |  | Male | 35-39 | 10 |  |
|  |  |  | Male | 40-44 | 12.5 |  |
|  |  |  | Male | 45-49 | 11.3 |  |
|  |  |  | Male | 10-49 | 6.8 |  |
|  |  |  | Female | 10-11 | 0.3 |  |
|  |  |  | Female | 12-14 | 0.8 |  |
|  |  |  | Female | 15-19 | 3.2 |  |
|  |  |  | Female | 20-24 | 6.6 |  |
|  |  |  | Female | 25-29 | 9.1 |  |
|  |  |  | Female | 30-34 | 12.7 |  |
|  |  |  | Female | 35-39 | 16.5 |  |
|  |  |  | Female | 40-44 | 20.5 |  |
|  |  |  | Female | 45-49 | 19.8 |  |
|  |  |  | Female | 10-49 | 11.3 |  |
|  |  | 2000 | Male | 10-11 | 0.7 |  |
|  |  |  | Male | 12-14 | 1.2 |  |
|  |  |  | Male | 15-19 | 3.3 |  |
|  |  |  | Male | 20-24 | 7.2 |  |
|  |  |  | Male | 25-29 | 8.7 |  |
|  |  |  | Male | 30-34 | 10.1 |  |
|  |  |  | Male | 35-39 | 10.8 |  |
|  |  |  | Male | 40-44 | 12.6 |  |
|  |  |  | Male | 45-49 | 11.3 |  |
|  |  |  | Male | 10-49 | 8.3 |  |
|  |  |  | Female | 10-11 | 0.4 |  |
|  |  |  | Female | 12-14 | 1.5 |  |
|  |  |  | Female | 15-19 | 8 |  |
|  |  |  | Female | 20-24 | 13.3 |  |
|  |  |  | Female | 25-29 | 14 |  |
|  |  |  | Female | 30-34 | 14.9 |  |
|  |  |  | Female | 35-39 | 17.4 |  |
|  |  |  | Female | 40-44 | 19 |  |
|  |  |  | Female | 45-49 | 17.8 |  |
|  |  |  | Female | 10-49 | 13.1 |  |
|  |  | 2005 | Male | 10-11 | 0.9 |  |
|  |  |  | Male | 12-14 | 1.8 |  |
|  |  |  | Male | 15-19 | 7 |  |
|  |  |  | Male | 20-24 | 10.9 |  |
|  |  |  | Male | 25-29 | 11.9 |  |
|  |  |  | Male | 30-34 | 11.9 |  |
|  |  |  | Male | 35-39 | 12.7 |  |
|  |  |  | Male | 40-44 | 13.6 |  |
|  |  |  | Male | 45-49 | 12.4 |  |
|  |  |  | Male | 10-49 | 10 |  |
|  |  |  | Female | 10-11 | 0.6 |  |
|  |  |  | Female | 12-14 | 3.4 |  |
|  |  |  | Female | 15-19 | 15.6 |  |
|  |  |  | Female | 20-24 | 20.9 |  |
|  |  |  | Female | 25-29 | 19.3 |  |
|  |  |  | Female | 30-34 | 19.4 |  |
|  |  |  | Female | 35-39 | 20.2 |  |
|  |  |  | Female | 40-44 | 20.8 |  |
|  |  |  | Female | 45-49 | 18.6 |  |
|  |  |  | Female | 10-49 | 16.5 |  |
|  |  | 2010 | Male | 10-11 | 1 |  |
|  |  |  | Male | 12-14 | 2.7 |  |
|  |  |  | Male | 15-19 | 11.1 |  |
|  |  |  | Male | 20-24 | 17.2 |  |
|  |  |  | Male | 25-29 | 16.6 |  |
|  |  |  | Male | 30-34 | 16.1 |  |
|  |  |  | Male | 35-39 | 15.2 |  |
|  |  |  | Male | 40-44 | 15.6 |  |
|  |  |  | Male | 45-49 | 13.8 |  |
|  |  |  | Male | 10-49 | 12.9 |  |
|  |  |  | Female | 10-11 | 1.1 |  |
|  |  |  | Female | 12-14 | 5.5 |  |
|  |  |  | Female | 15-19 | 25.2 |  |
|  |  |  | Female | 20-24 | 30.7 |  |
|  |  |  | Female | 25-29 | 26 |  |
|  |  |  | Female | 30-34 | 24.7 |  |
|  |  |  | Female | 35-39 | 23.3 |  |
|  |  |  | Female | 40-44 | 21.9 |  |
|  |  |  | Female | 45-49 | 19.3 |  |
|  |  |  | Female | 10-49 | 20.8 |  |
|  |  | 2013 | Male | 10-11 | 0.9 |  |
|  |  |  | Male | 12-14 | 1.8 |  |
|  |  |  | Male | 15-19 | 6.6 |  |
|  |  |  | Male | 20-24 | 11.7 |  |
|  |  |  | Male | 25-29 | 11.5 |  |
|  |  |  | Male | 30-34 | 10.5 |  |
|  |  |  | Male | 35-39 | 10 |  |
|  |  |  | Male | 40-44 | 10.1 |  |
|  |  |  | Male | 45-49 | 8.9 |  |
|  |  |  | Male | 10-49 | 8.4 |  |
|  |  |  | Female | 10-11 | 0.7 |  |
|  |  |  | Female | 12-14 | 3.5 |  |
|  |  |  | Female | 15-19 | 15.6 |  |
|  |  |  | Female | 20-24 | 17.9 |  |
|  |  |  | Female | 25-29 | 15 |  |
|  |  |  | Female | 30-34 | 15 |  |
|  |  |  | Female | 35-39 | 14.9 |  |
|  |  |  | Female | 40-44 | 13.3 |  |
|  |  |  | Female | 45-49 | 12.2 |  |
|  |  |  | Female | 10-49 | 12.7 |  |
| Rasmussen 2024^a^ [] | Denmark | 2007 | Combined | 10-13 | 1.14 | No previous dispensation since 2005 |
|  |  |  | Combined | 10-17 | 4.18 |  |
|  |  |  | Combined | 14-17 | 7.37 |  |
|  |  |  | Combined | 5-13 | 0.64 |  |
|  |  |  | Combined | 5-17 | 2.68 |  |
|  |  |  | Combined | 5-9 | 0.22 |  |
|  |  |  | Male | 10-13 | 1.19 |  |
|  |  |  | Male | 10-17 | 2.71 |  |
|  |  |  | Male | 14-17 | 4.31 |  |
|  |  |  | Male | 5-13 | 0.71 |  |
|  |  |  | Male | 5-17 | 1.81 |  |
|  |  |  | Male | 5-9 | 0.31 |  |
|  |  |  | Female | 10-13 | 1.09 |  |
|  |  |  | Female | 10-17 | 5.73 |  |
|  |  |  | Female | 14-17 | 10.60 |  |
|  |  |  | Female | 5-13 | 0.56 |  |
|  |  |  | Female | 5-17 | 3.60 |  |
|  |  |  | Female | 5-9 | 0.12 |  |
|  |  | 2008 | Combined | 10-13 | 1.17 |  |
|  |  |  | Combined | 10-17 | 4.48 |  |
|  |  |  | Combined | 14-17 | 7.88 |  |
|  |  |  | Combined | 5-13 | 0.66 |  |
|  |  |  | Combined | 5-17 | 2.90 |  |
|  |  |  | Combined | 5-9 | 0.23 |  |
|  |  |  | Male | 10-13 | 1.25 |  |
|  |  |  | Male | 10-17 | 2.75 |  |
|  |  |  | Male | 14-17 | 4.28 |  |
|  |  |  | Male | 5-13 | 0.76 |  |
|  |  |  | Male | 5-17 | 1.85 |  |
|  |  |  | Male | 5-9 | 0.33 |  |
|  |  |  | Female | 10-13 | 1.08 |  |
|  |  |  | Female | 10-17 | 6.32 |  |
|  |  |  | Female | 14-17 | 11.68 |  |
|  |  |  | Female | 5-13 | 0.56 |  |
|  |  |  | Female | 5-17 | 4.00 |  |
|  |  |  | Female | 5-9 | 0.13 |  |
|  |  | 2009 | Combined | 10-13 | 1.30 |  |
|  |  |  | Combined | 10-17 | 4.94 |  |
|  |  |  | Combined | 14-17 | 8.53 |  |
|  |  |  | Combined | 5-13 | 0.74 |  |
|  |  |  | Combined | 5-17 | 3.20 |  |
|  |  |  | Combined | 5-9 | 0.26 |  |
|  |  |  | Male | 10-13 | 1.33 |  |
|  |  |  | Male | 10-17 | 3.28 |  |
|  |  |  | Male | 14-17 | 5.22 |  |
|  |  |  | Male | 5-13 | 0.76 |  |
|  |  |  | Male | 5-17 | 2.17 |  |
|  |  |  | Male | 5-9 | 0.28 |  |
|  |  |  | Female | 10-13 | 1.28 |  |
|  |  |  | Female | 10-17 | 6.69 |  |
|  |  |  | Female | 14-17 | 12.02 |  |
|  |  |  | Female | 5-13 | 0.71 |  |
|  |  |  | Female | 5-17 | 4.29 |  |
|  |  |  | Female | 5-9 | 0.24 |  |
|  |  | 2010 | Combined | 10-13 | 1.42 |  |
|  |  |  | Combined | 10-17 | 5.58 |  |
|  |  |  | Combined | 14-17 | 9.55 |  |
|  |  |  | Combined | 5-13 | 0.79 |  |
|  |  |  | Combined | 5-17 | 3.61 |  |
|  |  |  | Combined | 5-9 | 0.28 |  |
|  |  |  | Male | 10-13 | 1.39 |  |
|  |  |  | Male | 10-17 | 3.36 |  |
|  |  |  | Male | 14-17 | 5.24 |  |
|  |  |  | Male | 5-13 | 0.82 |  |
|  |  |  | Male | 5-17 | 2.25 |  |
|  |  |  | Male | 5-9 | 0.36 |  |
|  |  |  | Female | 10-13 | 1.45 |  |
|  |  |  | Female | 10-17 | 7.91 |  |
|  |  |  | Female | 14-17 | 14.09 |  |
|  |  |  | Female | 5-13 | 0.76 |  |
|  |  |  | Female | 5-17 | 5.04 |  |
|  |  |  | Female | 5-9 | 0.20 |  |
|  |  | 2011 | Combined | 10-13 | 1.35 |  |
|  |  |  | Combined | 10-17 | 4.35 |  |
|  |  |  | Combined | 14-17 | 7.22 |  |
|  |  |  | Combined | 5-13 | 0.73 |  |
|  |  |  | Combined | 5-17 | 2.82 |  |
|  |  |  | Combined | 5-9 | 0.22 |  |
|  |  |  | Male | 10-13 | 1.42 |  |
|  |  |  | Male | 10-17 | 3.00 |  |
|  |  |  | Male | 14-17 | 4.51 |  |
|  |  |  | Male | 5-13 | 0.78 |  |
|  |  |  | Male | 5-17 | 1.98 |  |
|  |  |  | Male | 5-9 | 0.24 |  |
|  |  |  | Female | 10-13 | 1.26 |  |
|  |  |  | Female | 10-17 | 5.77 |  |
|  |  |  | Female | 14-17 | 10.08 |  |
|  |  |  | Female | 5-13 | 0.68 |  |
|  |  |  | Female | 5-17 | 3.71 |  |
|  |  |  | Female | 5-9 | 0.20 |  |
|  |  | 2012 | Combined | 10-13 | 1.23 |  |
|  |  |  | Combined | 10-17 | 4.08 |  |
|  |  |  | Combined | 14-17 | 6.76 |  |
|  |  |  | Combined | 5-13 | 0.64 |  |
|  |  |  | Combined | 5-17 | 2.62 |  |
|  |  |  | Combined | 5-9 | 0.16 |  |
|  |  |  | Male | 10-13 | 1.28 |  |
|  |  |  | Male | 10-17 | 2.55 |  |
|  |  |  | Male | 14-17 | 3.75 |  |
|  |  |  | Male | 5-13 | 0.70 |  |
|  |  |  | Male | 5-17 | 1.69 |  |
|  |  |  | Male | 5-9 | 0.23 |  |
|  |  |  | Female | 10-13 | 1.18 |  |
|  |  |  | Female | 10-17 | 5.68 |  |
|  |  |  | Female | 14-17 | 9.94 |  |
|  |  |  | Female | 5-13 | 0.58 |  |
|  |  |  | Female | 5-17 | 3.60 |  |
|  |  |  | Female | 5-9 | 0.09 |  |
|  |  | 2013 | Combined | 10-13 | 1.05 |  |
|  |  |  | Combined | 10-17 | 3.34 |  |
|  |  |  | Combined | 14-17 | 5.52 |  |
|  |  |  | Combined | 5-13 | 0.53 |  |
|  |  |  | Combined | 5-17 | 2.13 |  |
|  |  |  | Combined | 5-9 | 0.12 |  |
|  |  |  | Male | 10-13 | 1.14 |  |
|  |  |  | Male | 10-17 | 2.05 |  |
|  |  |  | Male | 14-17 | 2.91 |  |
|  |  |  | Male | 5-13 | 0.58 |  |
|  |  |  | Male | 5-17 | 1.32 |  |
|  |  |  | Male | 5-9 | 0.12 |  |
|  |  |  | Female | 10-13 | 0.95 |  |
|  |  |  | Female | 10-17 | 4.69 |  |
|  |  |  | Female | 14-17 | 8.27 |  |
|  |  |  | Female | 5-13 | 0.49 |  |
|  |  |  | Female | 5-17 | 2.97 |  |
|  |  |  | Female | 5-9 | 0.11 |  |
|  |  | 2014 | Combined | 10-13 | 1.02 |  |
|  |  |  | Combined | 10-17 | 2.92 |  |
|  |  |  | Combined | 14-17 | 4.74 |  |
|  |  |  | Combined | 5-13 | 0.51 |  |
|  |  |  | Combined | 5-17 | 1.85 |  |
|  |  |  | Combined | 5-9 | 0.11 |  |
|  |  |  | Male | 10-13 | 1.13 |  |
|  |  |  | Male | 10-17 | 1.84 |  |
|  |  |  | Male | 14-17 | 2.53 |  |
|  |  |  | Male | 5-13 | 0.59 |  |
|  |  |  | Male | 5-17 | 1.20 |  |
|  |  |  | Male | 5-9 | 0.16 |  |
|  |  |  | Female | 10-13 | 0.90 |  |
|  |  |  | Female | 10-17 | 4.04 |  |
|  |  |  | Female | 14-17 | 7.07 |  |
|  |  |  | Female | 5-13 | 0.43 |  |
|  |  |  | Female | 5-17 | 2.53 |  |
|  |  |  | Female | 5-9 | 0.06 |  |
|  |  | 2015 | Combined | 10-13 | 0.93 |  |
|  |  |  | Combined | 10-17 | 2.88 |  |
|  |  |  | Combined | 14-17 | 4.74 |  |
|  |  |  | Combined | 5-13 | 0.47 |  |
|  |  |  | Combined | 5-17 | 1.82 |  |
|  |  |  | Combined | 5-9 | 0.10 |  |
|  |  |  | Male | 10-13 | 1.03 |  |
|  |  |  | Male | 10-17 | 1.87 |  |
|  |  |  | Male | 14-17 | 2.67 |  |
|  |  |  | Male | 5-13 | 0.52 |  |
|  |  |  | Male | 5-17 | 1.20 |  |
|  |  |  | Male | 5-9 | 0.12 |  |
|  |  |  | Female | 10-13 | 0.84 |  |
|  |  |  | Female | 10-17 | 3.94 |  |
|  |  |  | Female | 14-17 | 6.91 |  |
|  |  |  | Female | 5-13 | 0.42 |  |
|  |  |  | Female | 5-17 | 2.47 |  |
|  |  |  | Female | 5-9 | 0.08 |  |
|  |  | 2016 | Combined | 10-13 | 0.83 |  |
|  |  |  | Combined | 10-17 | 2.62 |  |
|  |  |  | Combined | 14-17 | 4.35 |  |
|  |  |  | Combined | 5-13 | 0.42 |  |
|  |  |  | Combined | 5-17 | 1.66 |  |
|  |  |  | Combined | 5-9 | 0.10 |  |
|  |  |  | Male | 10-13 | 0.86 |  |
|  |  |  | Male | 10-17 | 1.65 |  |
|  |  |  | Male | 14-17 | 2.42 |  |
|  |  |  | Male | 5-13 | 0.45 |  |
|  |  |  | Male | 5-17 | 1.07 |  |
|  |  |  | Male | 5-9 | 0.12 |  |
|  |  |  | Female | 10-13 | 0.80 |  |
|  |  |  | Female | 10-17 | 3.64 |  |
|  |  |  | Female | 14-17 | 6.38 |  |
|  |  |  | Female | 5-13 | 0.39 |  |
|  |  |  | Female | 5-17 | 2.28 |  |
|  |  |  | Female | 5-9 | 0.07 |  |
|  |  | 2017 | Combined | 10-13 | 0.88 |  |
|  |  |  | Combined | 10-17 | 2.48 |  |
|  |  |  | Combined | 14-17 | 4.05 |  |
|  |  |  | Combined | 5-13 | 0.43 |  |
|  |  |  | Combined | 5-17 | 1.57 |  |
|  |  |  | Combined | 5-9 | 0.07 |  |
|  |  |  | Male | 10-13 | 0.80 |  |
|  |  |  | Male | 10-17 | 1.58 |  |
|  |  |  | Male | 14-17 | 2.36 |  |
|  |  |  | Male | 5-13 | 0.41 |  |
|  |  |  | Male | 5-17 | 1.02 |  |
|  |  |  | Male | 5-9 | 0.08 |  |
|  |  |  | Female | 10-13 | 0.97 |  |
|  |  |  | Female | 10-17 | 3.41 |  |
|  |  |  | Female | 14-17 | 5.82 |  |
|  |  |  | Female | 5-13 | 0.46 |  |
|  |  |  | Female | 5-17 | 2.14 |  |
|  |  |  | Female | 5-9 | 0.05 |  |
|  |  | 2018 | Combined | 10-13 | 0.91 |  |
|  |  |  | Combined | 10-17 | 2.68 |  |
|  |  |  | Combined | 14-17 | 4.43 |  |
|  |  |  | Combined | 5-13 | 0.45 |  |
|  |  |  | Combined | 5-17 | 1.70 |  |
|  |  |  | Combined | 5-9 | 0.07 |  |
|  |  |  | Male | 10-13 | 0.85 |  |
|  |  |  | Male | 10-17 | 1.82 |  |
|  |  |  | Male | 14-17 | 2.78 |  |
|  |  |  | Male | 5-13 | 0.44 |  |
|  |  |  | Male | 5-17 | 1.18 |  |
|  |  |  | Male | 5-9 | 0.10 |  |
|  |  |  | Female | 10-13 | 0.98 |  |
|  |  |  | Female | 10-17 | 3.57 |  |
|  |  |  | Female | 14-17 | 6.15 |  |
|  |  |  | Female | 5-13 | 0.47 |  |
|  |  |  | Female | 5-17 | 2.26 |  |
|  |  |  | Female | 5-9 | 0.04 |  |
| Ishtiak-Ahmed 2023^a^ [] | Denmark | 2016 | Combined | 65 and older | 32.3 | No dispensation in the previous 1 year |
|  |  |  | Male | 65 and older | 27.4 |  |
|  |  |  | Female | 65 and older | 36.8 |  |
|  |  | 2017 | Combined | 65 and older | 30.2 |  |
|  |  |  | Male | 65 and older | 25.5 |  |
|  |  |  | Female | 65 and older | 34.5 |  |
|  |  | 2018 | Combined | 65 and older | 29.7 |  |
|  |  |  | Male | 65 and older | 25.7 |  |
|  |  |  | Female | 65 and older | 33.4 |  |
| Saastamoinen 2012^a^ [] | Finland | 1999 | Combined | 0-17 | 1.06 | No dispensation in the previous 1 year |
|  |  |  | Combined | 0-11 | 0.15 |  |
|  |  |  | Combined | 12-15 | 1.69 |  |
|  |  |  | Combined | 16-17 | 4.96 |  |
|  |  | 2000 | Combined | 0-17 | 1.64 |  |
|  |  |  | Combined | 0-11 | 0.22 |  |
|  |  |  | Combined | 12-15 | 2.42 |  |
|  |  |  | Combined | 16-17 | 8.07 |  |
|  |  | 2001 | Combined | 0-17 | 1.51 |  |
|  |  |  | Combined | 0-11 | 0.20 |  |
|  |  |  | Combined | 12-15 | 2.37 |  |
|  |  |  | Combined | 16-17 | 7.24 |  |
|  |  | 2002 | Combined | 0-17 | 1.71 |  |
|  |  |  | Combined | 0-11 | 0.27 |  |
|  |  |  | Combined | 12-15 | 2.71 |  |
|  |  |  | Combined | 16-17 | 8.01 |  |
|  |  | 2003 | Combined | 0-17 | 1.78 |  |
|  |  |  | Combined | 0-11 | 0.33 |  |
|  |  |  | Combined | 12-15 | 2.91 |  |
|  |  |  | Combined | 16-17 | 7.89 |  |
|  |  | 2004 | Combined | 0-17 | 1.56 |  |
|  |  |  | Combined | 0-11 | 0.26 |  |
|  |  |  | Combined | 12-15 | 2.61 |  |
|  |  |  | Combined | 16-17 | 6.72 |  |
|  |  | 2005 | Combined | 0-17 | 1.68 |  |
|  |  |  | Combined | 0-11 | 0.23 |  |
|  |  |  | Combined | 12-15 | 2.73 |  |
|  |  |  | Combined | 16-17 | 7.38 |  |
| Foulon 2010^b^ [] | Finland | 1999 | Combined | 0-19 | 2.01 | No dispensation in the previous 6 months |
|  |  | 2002 | Combined | 0-19 | 3.02 |  |
|  |  | 2005 | Combined | 0-19 | 3.12 |  |
| Autti-Rämö 2011^a^ [] | Finland | 1997 | Combined | 0-26 | 4.00 | No dispensation in the previous 1 year |
|  |  |  | Combined | 0-6 | 0.00 |  |
|  |  |  | Combined | 7-10 | 0.40 |  |
|  |  |  | Combined | 11-15 | 1.10 |  |
|  |  |  | Combined | 16-20 | 5.70 |  |
|  |  |  | Combined | 21-26 | 12.50 |  |
|  |  |  | Male | 0-26 | 3.30 |  |
|  |  |  | Male | 0-6 | 0.10 |  |
|  |  |  | Male | 7-10 | 0.40 |  |
|  |  |  | Male | 11-15 | 1.00 |  |
|  |  |  | Male | 16-20 | 3.70 |  |
|  |  |  | Male | 21-26 | 10.80 |  |
|  |  |  | Female | 0-26 | 4.80 |  |
|  |  |  | Female | 0-6 | 0.00 |  |
|  |  |  | Female | 7-10 | 0.30 |  |
|  |  |  | Female | 11-15 | 1.10 |  |
|  |  |  | Female | 16-20 | 7.80 |  |
|  |  |  | Female | 21-26 | 14.20 |  |
|  |  | 2002 | Combined | 0-26 | 7.70 |  |
|  |  |  | Combined | 0-6 | 0.00 |  |
|  |  |  | Combined | 7-10 | 0.50 |  |
|  |  |  | Combined | 11-15 | 2.70 |  |
|  |  |  | Combined | 16-20 | 13.20 |  |
|  |  |  | Combined | 21-26 | 19.90 |  |
|  |  |  | Male | 0-26 | 6.00 |  |
|  |  |  | Male | 0-6 | 0.10 |  |
|  |  |  | Male | 7-10 | 0.80 |  |
|  |  |  | Male | 11-15 | 2.20 |  |
|  |  |  | Male | 16-20 | 8.80 |  |
|  |  |  | Male | 21-26 | 16.50 |  |
|  |  |  | Female | 0-26 | 9.50 |  |
|  |  |  | Female | 0-6 | 0.00 |  |
|  |  |  | Female | 7-10 | 0.20 |  |
|  |  |  | Female | 11-15 | 3.20 |  |
|  |  |  | Female | 16-20 | 17.90 |  |
|  |  |  | Female | 21-26 | 23.50 |  |
|  |  | 2007 | Combined | 0-26 | 11.50 |  |
|  |  |  | Combined | 0-6 | 0.00 |  |
|  |  |  | Combined | 7-10 | 0.60 |  |
|  |  |  | Combined | 11-15 | 3.40 |  |
|  |  |  | Combined | 16-20 | 18.70 |  |
|  |  |  | Combined | 21-26 | 30.20 |  |
|  |  |  | Male | 0-26 | 8.50 |  |
|  |  |  | Male | 0-6 | 0.00 |  |
|  |  |  | Male | 7-10 | 0.80 |  |
|  |  |  | Male | 11-15 | 2.60 |  |
|  |  |  | Male | 16-20 | 11.70 |  |
|  |  |  | Male | 21-26 | 23.80 |  |
|  |  |  | Female | 0-26 | 14.60 |  |
|  |  |  | Female | 0-6 | 0.00 |  |
|  |  |  | Female | 7-10 | 0.30 |  |
|  |  |  | Female | 11-15 | 4.20 |  |
|  |  |  | Female | 16-20 | 26.00 |  |
|  |  |  | Female | 21-26 | 36.90 |  |
| Zoëga 2009^a^ [] | Iceland | 2004 | Combined | 0-17 | 11.90 | No dispensation in the previous 1 year |
|  |  |  | Combined | 0-5 | 3.70 |  |
|  |  |  | Combined | 6-11 | 12.50 |  |
|  |  |  | Combined | 12-17 | 18.90 |  |
|  |  |  | Male | 0-17 | 12.60 |  |
|  |  |  | Female | 0-17 | 11.10 |  |
|  |  | 2005 | Combined | 0-17 | 9.80 |  |
|  |  |  | Combined | 0-5 | 2.40 |  |
|  |  |  | Combined | 6-11 | 9.10 |  |
|  |  |  | Combined | 12-17 | 17.20 |  |
|  |  |  | Male | 0-17 | 9.90 |  |
|  |  |  | Female | 0-17 | 9.60 |  |
|  |  | 2006 | Combined | 0-17 | 8.40 |  |
|  |  |  | Combined | 0-5 | 2.10 |  |
|  |  |  | Combined | 6-11 | 8.80 |  |
|  |  |  | Combined | 12-17 | 13.70 |  |
|  |  |  | Male | 0-17 | 8.50 |  |
|  |  |  | Female | 0-17 | 8.20 |  |
|  |  | 2007 | Combined | 0-17 | 8.00 |  |
|  |  |  | Combined | 0-5 | 1.60 |  |
|  |  |  | Combined | 6-11 | 8.70 |  |
|  |  |  | Combined | 12-17 | 13.40 |  |
|  |  |  | Male | 0-17 | 8.30 |  |
|  |  |  | Female | 0-17 | 7.80 |  |
| Kjosavik 2011^b^ [] | Norway | 2008 | Combined | 0-19 | 1.5 | No dispensation in the previous 3 years |
|  |  |  | Combined | 20-39 | 10.5 |  |
|  |  |  | Combined | 40-59 | 11.3 |  |
|  |  |  | Combined | 60-69 | 10.3 |  |
|  |  |  | Combined | 70-79 | 13.6 |  |
|  |  |  | Combined | 80 and older | 18.0 |  |
|  |  |  | Male | 0-19 | 1 |  |
|  |  |  | Male | 20-39 | 8.5 |  |
|  |  |  | Male | 40-59 | 8.8 |  |
|  |  |  | Male | 60-69 | 8.4 |  |
|  |  |  | Male | 70-79 | 10.6 |  |
|  |  |  | Male | 80 and older | 15.6 |  |
|  |  |  | Female | 0-19 | 2 |  |
|  |  |  | Female | 20-39 | 12.7 |  |
|  |  |  | Female | 40-59 | 14.1 |  |
|  |  |  | Female | 60-69 | 12.4 |  |
|  |  |  | Female | 70-79 | 16.2 |  |
|  |  |  | Female | 80 and older | 19.5 |  |
| Rasmussen 2024^a^ [] | Norway | 2007 | Combined | 10-13 | 0.75 | No previous dispensation since 2005 |
|  |  |  | Combined | 10-17 | 3.13 |  |
|  |  |  | Combined | 14-17 | 5.48 |  |
|  |  |  | Combined | 5-13 | 0.44 |  |
|  |  |  | Combined | 5-17 | 2.02 |  |
|  |  |  | Combined | 5-9 | 0.18 |  |
|  |  |  | Male | 10-13 | 0.95 |  |
|  |  |  | Male | 10-17 | 2.17 |  |
|  |  |  | Male | 14-17 | 3.38 |  |
|  |  |  | Male | 5-13 | 0.57 |  |
|  |  |  | Male | 5-17 | 1.45 |  |
|  |  |  | Male | 5-9 | 0.26 |  |
|  |  |  | Female | 10-13 | 0.54 |  |
|  |  |  | Female | 10-17 | 4.14 |  |
|  |  |  | Female | 14-17 | 7.71 |  |
|  |  |  | Female | 5-13 | 0.30 |  |
|  |  |  | Female | 5-17 | 2.62 |  |
|  |  |  | Female | 5-9 | 0.10 |  |
|  |  | 2008 | Combined | 10-13 | 0.80 |  |
|  |  |  | Combined | 10-17 | 3.20 |  |
|  |  |  | Combined | 14-17 | 5.58 |  |
|  |  |  | Combined | 5-13 | 0.47 |  |
|  |  |  | Combined | 5-17 | 2.08 |  |
|  |  |  | Combined | 5-9 | 0.18 |  |
|  |  |  | Male | 10-13 | 0.99 |  |
|  |  |  | Male | 10-17 | 2.14 |  |
|  |  |  | Male | 14-17 | 3.29 |  |
|  |  |  | Male | 5-13 | 0.60 |  |
|  |  |  | Male | 5-17 | 1.45 |  |
|  |  |  | Male | 5-9 | 0.27 |  |
|  |  |  | Female | 10-13 | 0.60 |  |
|  |  |  | Female | 10-17 | 4.32 |  |
|  |  |  | Female | 14-17 | 7.99 |  |
|  |  |  | Female | 5-13 | 0.33 |  |
|  |  |  | Female | 5-17 | 2.75 |  |
|  |  |  | Female | 5-9 | 0.10 |  |
|  |  | 2009 | Combined | 10-13 | 0.93 |  |
|  |  |  | Combined | 10-17 | 3.17 |  |
|  |  |  | Combined | 14-17 | 5.38 |  |
|  |  |  | Combined | 5-13 | 0.51 |  |
|  |  |  | Combined | 5-17 | 2.05 |  |
|  |  |  | Combined | 5-9 | 0.15 |  |
|  |  |  | Male | 10-13 | 1.05 |  |
|  |  |  | Male | 10-17 | 2.29 |  |
|  |  |  | Male | 14-17 | 3.51 |  |
|  |  |  | Male | 5-13 | 0.59 |  |
|  |  |  | Male | 5-17 | 1.51 |  |
|  |  |  | Male | 5-9 | 0.20 |  |
|  |  |  | Female | 10-13 | 0.81 |  |
|  |  |  | Female | 10-17 | 4.10 |  |
|  |  |  | Female | 14-17 | 7.35 |  |
|  |  |  | Female | 5-13 | 0.42 |  |
|  |  |  | Female | 5-17 | 2.61 |  |
|  |  |  | Female | 5-9 | 0.10 |  |
|  |  | 2010 | Combined | 10-13 | 0.89 |  |
|  |  |  | Combined | 10-17 | 3.32 |  |
|  |  |  | Combined | 14-17 | 5.72 |  |
|  |  |  | Combined | 5-13 | 0.52 |  |
|  |  |  | Combined | 5-17 | 2.17 |  |
|  |  |  | Combined | 5-9 | 0.21 |  |
|  |  |  | Male | 10-13 | 0.88 |  |
|  |  |  | Male | 10-17 | 2.40 |  |
|  |  |  | Male | 14-17 | 3.88 |  |
|  |  |  | Male | 5-13 | 0.56 |  |
|  |  |  | Male | 5-17 | 1.62 |  |
|  |  |  | Male | 5-9 | 0.28 |  |
|  |  |  | Female | 10-13 | 0.89 |  |
|  |  |  | Female | 10-17 | 4.30 |  |
|  |  |  | Female | 14-17 | 7.67 |  |
|  |  |  | Female | 5-13 | 0.48 |  |
|  |  |  | Female | 5-17 | 2.75 |  |
|  |  |  | Female | 5-9 | 0.14 |  |
|  |  | 2011 | Combined | 10-13 | 0.88 |  |
|  |  |  | Combined | 10-17 | 3.56 |  |
|  |  |  | Combined | 14-17 | 6.17 |  |
|  |  |  | Combined | 5-13 | 0.50 |  |
|  |  |  | Combined | 5-17 | 2.31 |  |
|  |  |  | Combined | 5-9 | 0.18 |  |
|  |  |  | Male | 10-13 | 1.08 |  |
|  |  |  | Male | 10-17 | 2.32 |  |
|  |  |  | Male | 14-17 | 3.52 |  |
|  |  |  | Male | 5-13 | 0.61 |  |
|  |  |  | Male | 5-17 | 1.55 |  |
|  |  |  | Male | 5-9 | 0.22 |  |
|  |  |  | Female | 10-13 | 0.68 |  |
|  |  |  | Female | 10-17 | 4.88 |  |
|  |  |  | Female | 14-17 | 8.98 |  |
|  |  |  | Female | 5-13 | 0.38 |  |
|  |  |  | Female | 5-17 | 3.12 |  |
|  |  |  | Female | 5-9 | 0.13 |  |
|  |  | 2012 | Combined | 10-13 | 1.03 |  |
|  |  |  | Combined | 10-17 | 4.21 |  |
|  |  |  | Combined | 14-17 | 7.25 |  |
|  |  |  | Combined | 5-13 | 0.54 |  |
|  |  |  | Combined | 5-17 | 2.69 |  |
|  |  |  | Combined | 5-9 | 0.14 |  |
|  |  |  | Male | 10-13 | 1.19 |  |
|  |  |  | Male | 10-17 | 2.69 |  |
|  |  |  | Male | 14-17 | 4.11 |  |
|  |  |  | Male | 5-13 | 0.64 |  |
|  |  |  | Male | 5-17 | 1.76 |  |
|  |  |  | Male | 5-9 | 0.18 |  |
|  |  |  | Female | 10-13 | 0.87 |  |
|  |  |  | Female | 10-17 | 5.82 |  |
|  |  |  | Female | 14-17 | 10.59 |  |
|  |  |  | Female | 5-13 | 0.45 |  |
|  |  |  | Female | 5-17 | 3.68 |  |
|  |  |  | Female | 5-9 | 0.10 |  |
|  |  | 2013 | Combined | 10-13 | 0.88 |  |
|  |  |  | Combined | 10-17 | 4.29 |  |
|  |  |  | Combined | 14-17 | 7.54 |  |
|  |  |  | Combined | 5-13 | 0.49 |  |
|  |  |  | Combined | 5-17 | 2.73 |  |
|  |  |  | Combined | 5-9 | 0.17 |  |
|  |  |  | Male | 10-13 | 0.89 |  |
|  |  |  | Male | 10-17 | 2.70 |  |
|  |  |  | Male | 14-17 | 4.42 |  |
|  |  |  | Male | 5-13 | 0.53 |  |
|  |  |  | Male | 5-17 | 1.77 |  |
|  |  |  | Male | 5-9 | 0.24 |  |
|  |  |  | Female | 10-13 | 0.87 |  |
|  |  |  | Female | 10-17 | 5.96 |  |
|  |  |  | Female | 14-17 | 10.84 |  |
|  |  |  | Female | 5-13 | 0.44 |  |
|  |  |  | Female | 5-17 | 3.74 |  |
|  |  |  | Female | 5-9 | 0.09 |  |
|  |  | 2014 | Combined | 10-13 | 0.92 |  |
|  |  |  | Combined | 10-17 | 4.34 |  |
|  |  |  | Combined | 14-17 | 7.57 |  |
|  |  |  | Combined | 5-13 | 0.48 |  |
|  |  |  | Combined | 5-17 | 2.73 |  |
|  |  |  | Combined | 5-9 | 0.14 |  |
|  |  |  | Male | 10-13 | 0.95 |  |
|  |  |  | Male | 10-17 | 2.61 |  |
|  |  |  | Male | 14-17 | 4.17 |  |
|  |  |  | Male | 5-13 | 0.50 |  |
|  |  |  | Male | 5-17 | 1.67 |  |
|  |  |  | Male | 5-9 | 0.15 |  |
|  |  |  | Female | 10-13 | 0.89 |  |
|  |  |  | Female | 10-17 | 6.15 |  |
|  |  |  | Female | 14-17 | 11.18 |  |
|  |  |  | Female | 5-13 | 0.46 |  |
|  |  |  | Female | 5-17 | 3.85 |  |
|  |  |  | Female | 5-9 | 0.13 |  |
|  |  | 2015 | Combined | 10-13 | 0.95 |  |
|  |  |  | Combined | 10-17 | 4.50 |  |
|  |  |  | Combined | 14-17 | 7.87 |  |
|  |  |  | Combined | 5-13 | 0.50 |  |
|  |  |  | Combined | 5-17 | 2.81 |  |
|  |  |  | Combined | 5-9 | 0.15 |  |
|  |  |  | Male | 10-13 | 1.04 |  |
|  |  |  | Male | 10-17 | 2.63 |  |
|  |  |  | Male | 14-17 | 4.13 |  |
|  |  |  | Male | 5-13 | 0.57 |  |
|  |  |  | Male | 5-17 | 1.69 |  |
|  |  |  | Male | 5-9 | 0.21 |  |
|  |  |  | Female | 10-13 | 0.85 |  |
|  |  |  | Female | 10-17 | 6.45 |  |
|  |  |  | Female | 14-17 | 11.81 |  |
|  |  |  | Female | 5-13 | 0.42 |  |
|  |  |  | Female | 5-17 | 3.99 |  |
|  |  |  | Female | 5-9 | 0.08 |  |
|  |  | 2016 | Combined | 10-13 | 0.89 |  |
|  |  |  | Combined | 10-17 | 4.42 |  |
|  |  |  | Combined | 14-17 | 7.82 |  |
|  |  |  | Combined | 5-13 | 0.45 |  |
|  |  |  | Combined | 5-17 | 2.74 |  |
|  |  |  | Combined | 5-9 | 0.12 |  |
|  |  |  | Male | 10-13 | 0.92 |  |
|  |  |  | Male | 10-17 | 2.61 |  |
|  |  |  | Male | 14-17 | 4.24 |  |
|  |  |  | Male | 5-13 | 0.48 |  |
|  |  |  | Male | 5-17 | 1.65 |  |
|  |  |  | Male | 5-9 | 0.15 |  |
|  |  |  | Female | 10-13 | 0.86 |  |
|  |  |  | Female | 10-17 | 6.31 |  |
|  |  |  | Female | 14-17 | 11.60 |  |
|  |  |  | Female | 5-13 | 0.42 |  |
|  |  |  | Female | 5-17 | 3.88 |  |
|  |  |  | Female | 5-9 | 0.09 |  |
|  |  | 2017 | Combined | 10-13 | 0.89 |  |
|  |  |  | Combined | 10-17 | 4.27 |  |
|  |  |  | Combined | 14-17 | 7.59 |  |
|  |  |  | Combined | 5-13 | 0.47 |  |
|  |  |  | Combined | 5-17 | 2.66 |  |
|  |  |  | Combined | 5-9 | 0.14 |  |
|  |  |  | Male | 10-13 | 0.81 |  |
|  |  |  | Male | 10-17 | 2.58 |  |
|  |  |  | Male | 14-17 | 4.31 |  |
|  |  |  | Male | 5-13 | 0.45 |  |
|  |  |  | Male | 5-17 | 1.64 |  |
|  |  |  | Male | 5-9 | 0.17 |  |
|  |  |  | Female | 10-13 | 0.96 |  |
|  |  |  | Female | 10-17 | 6.04 |  |
|  |  |  | Female | 14-17 | 11.07 |  |
|  |  |  | Female | 5-13 | 0.48 |  |
|  |  |  | Female | 5-17 | 3.73 |  |
|  |  |  | Female | 5-9 | 0.11 |  |
|  |  | 2018 | Combined | 10-13 | 0.87 |  |
|  |  |  | Combined | 10-17 | 3.97 |  |
|  |  |  | Combined | 14-17 | 7.09 |  |
|  |  |  | Combined | 5-13 | 0.46 |  |
|  |  |  | Combined | 5-17 | 2.48 |  |
|  |  |  | Combined | 5-9 | 0.13 |  |
|  |  |  | Male | 10-13 | 0.83 |  |
|  |  |  | Male | 10-17 | 2.61 |  |
|  |  |  | Male | 14-17 | 4.40 |  |
|  |  |  | Male | 5-13 | 0.45 |  |
|  |  |  | Male | 5-17 | 1.65 |  |
|  |  |  | Male | 5-9 | 0.15 |  |
|  |  |  | Female | 10-13 | 0.92 |  |
|  |  |  | Female | 10-17 | 5.41 |  |
|  |  |  | Female | 14-17 | 9.93 |  |
|  |  |  | Female | 5-13 | 0.47 |  |
|  |  |  | Female | 5-17 | 3.35 |  |
|  |  |  | Female | 5-9 | 0.11 |  |
| Rasmussen 2024^a^ [] | Sweden | 2007 | Combined | 10-13 | 1.71 | No previous dispensation since 2005 |
|  |  |  | Combined | 10-17 | 4.40 |  |
|  |  |  | Combined | 14-17 | 6.61 |  |
|  |  |  | Combined | 5-13 | 0.92 |  |
|  |  |  | Combined | 5-17 | 2.99 |  |
|  |  |  | Combined | 5-9 | 0.23 |  |
|  |  |  | Male | 10-13 | 1.78 |  |
|  |  |  | Male | 10-17 | 3.05 |  |
|  |  |  | Male | 14-17 | 4.08 |  |
|  |  |  | Male | 5-13 | 0.98 |  |
|  |  |  | Male | 5-17 | 2.11 |  |
|  |  |  | Male | 5-9 | 0.29 |  |
|  |  |  | Female | 10-13 | 1.63 |  |
|  |  |  | Female | 10-17 | 5.83 |  |
|  |  |  | Female | 14-17 | 9.28 |  |
|  |  |  | Female | 5-13 | 0.86 |  |
|  |  |  | Female | 5-17 | 3.91 |  |
|  |  |  | Female | 5-9 | 0.17 |  |
|  |  | 2008 | Combined | 10-13 | 1.88 |  |
|  |  |  | Combined | 10-17 | 4.45 |  |
|  |  |  | Combined | 14-17 | 6.49 |  |
|  |  |  | Combined | 5-13 | 0.98 |  |
|  |  |  | Combined | 5-17 | 2.97 |  |
|  |  |  | Combined | 5-9 | 0.24 |  |
|  |  |  | Male | 10-13 | 1.97 |  |
|  |  |  | Male | 10-17 | 3.18 |  |
|  |  |  | Male | 14-17 | 4.16 |  |
|  |  |  | Male | 5-13 | 1.05 |  |
|  |  |  | Male | 5-17 | 2.17 |  |
|  |  |  | Male | 5-9 | 0.30 |  |
|  |  |  | Female | 10-13 | 1.80 |  |
|  |  |  | Female | 10-17 | 5.78 |  |
|  |  |  | Female | 14-17 | 8.96 |  |
|  |  |  | Female | 5-13 | 0.91 |  |
|  |  |  | Female | 5-17 | 3.81 |  |
|  |  |  | Female | 5-9 | 0.18 |  |
|  |  | 2009 | Combined | 10-13 | 1.69 |  |
|  |  |  | Combined | 10-17 | 4.54 |  |
|  |  |  | Combined | 14-17 | 6.84 |  |
|  |  |  | Combined | 5-13 | 0.87 |  |
|  |  |  | Combined | 5-17 | 2.96 |  |
|  |  |  | Combined | 5-9 | 0.24 |  |
|  |  |  | Male | 10-13 | 1.77 |  |
|  |  |  | Male | 10-17 | 3.27 |  |
|  |  |  | Male | 14-17 | 4.48 |  |
|  |  |  | Male | 5-13 | 0.97 |  |
|  |  |  | Male | 5-17 | 2.20 |  |
|  |  |  | Male | 5-9 | 0.36 |  |
|  |  |  | Female | 10-13 | 1.59 |  |
|  |  |  | Female | 10-17 | 5.87 |  |
|  |  |  | Female | 14-17 | 9.32 |  |
|  |  |  | Female | 5-13 | 0.76 |  |
|  |  |  | Female | 5-17 | 3.76 |  |
|  |  |  | Female | 5-9 | 0.11 |  |
|  |  | 2010 | Combined | 10-13 | 2.14 |  |
|  |  |  | Combined | 10-17 | 5.09 |  |
|  |  |  | Combined | 14-17 | 7.59 |  |
|  |  |  | Combined | 5-13 | 1.11 |  |
|  |  |  | Combined | 5-17 | 3.29 |  |
|  |  |  | Combined | 5-9 | 0.34 |  |
|  |  |  | Male | 10-13 | 2.23 |  |
|  |  |  | Male | 10-17 | 3.80 |  |
|  |  |  | Male | 14-17 | 5.13 |  |
|  |  |  | Male | 5-13 | 1.21 |  |
|  |  |  | Male | 5-17 | 2.53 |  |
|  |  |  | Male | 5-9 | 0.44 |  |
|  |  |  | Female | 10-13 | 2.04 |  |
|  |  |  | Female | 10-17 | 6.46 |  |
|  |  |  | Female | 14-17 | 10.18 |  |
|  |  |  | Female | 5-13 | 1.01 |  |
|  |  |  | Female | 5-17 | 4.09 |  |
|  |  |  | Female | 5-9 | 0.23 |  |
|  |  | 2011 | Combined | 10-13 | 2.38 |  |
|  |  |  | Combined | 10-17 | 5.66 |  |
|  |  |  | Combined | 14-17 | 8.60 |  |
|  |  |  | Combined | 5-13 | 1.22 |  |
|  |  |  | Combined | 5-17 | 3.58 |  |
|  |  |  | Combined | 5-9 | 0.38 |  |
|  |  |  | Male | 10-13 | 2.40 |  |
|  |  |  | Male | 10-17 | 4.25 |  |
|  |  |  | Male | 14-17 | 5.91 |  |
|  |  |  | Male | 5-13 | 1.29 |  |
|  |  |  | Male | 5-17 | 2.77 |  |
|  |  |  | Male | 5-9 | 0.48 |  |
|  |  |  | Female | 10-13 | 2.35 |  |
|  |  |  | Female | 10-17 | 7.15 |  |
|  |  |  | Female | 14-17 | 11.45 |  |
|  |  |  | Female | 5-13 | 1.15 |  |
|  |  |  | Female | 5-17 | 4.44 |  |
|  |  |  | Female | 5-9 | 0.27 |  |
|  |  | 2012 | Combined | 10-13 | 2.61 |  |
|  |  |  | Combined | 10-17 | 6.10 |  |
|  |  |  | Combined | 14-17 | 9.45 |  |
|  |  |  | Combined | 5-13 | 1.37 |  |
|  |  |  | Combined | 5-17 | 3.84 |  |
|  |  |  | Combined | 5-9 | 0.47 |  |
|  |  |  | Male | 10-13 | 2.73 |  |
|  |  |  | Male | 10-17 | 4.59 |  |
|  |  |  | Male | 14-17 | 6.36 |  |
|  |  |  | Male | 5-13 | 1.52 |  |
|  |  |  | Male | 5-17 | 3.00 |  |
|  |  |  | Male | 5-9 | 0.64 |  |
|  |  |  | Female | 10-13 | 2.49 |  |
|  |  |  | Female | 10-17 | 7.71 |  |
|  |  |  | Female | 14-17 | 12.75 |  |
|  |  |  | Female | 5-13 | 1.21 |  |
|  |  |  | Female | 5-17 | 4.72 |  |
|  |  |  | Female | 5-9 | 0.29 |  |
|  |  | 2013 | Combined | 10-13 | 2.82 |  |
|  |  |  | Combined | 10-17 | 6.58 |  |
|  |  |  | Combined | 14-17 | 10.42 |  |
|  |  |  | Combined | 5-13 | 1.44 |  |
|  |  |  | Combined | 5-17 | 4.08 |  |
|  |  |  | Combined | 5-9 | 0.43 |  |
|  |  |  | Male | 10-13 | 2.90 |  |
|  |  |  | Male | 10-17 | 4.77 |  |
|  |  |  | Male | 14-17 | 6.67 |  |
|  |  |  | Male | 5-13 | 1.57 |  |
|  |  |  | Male | 5-17 | 3.08 |  |
|  |  |  | Male | 5-9 | 0.59 |  |
|  |  |  | Female | 10-13 | 2.73 |  |
|  |  |  | Female | 10-17 | 8.50 |  |
|  |  |  | Female | 14-17 | 14.41 |  |
|  |  |  | Female | 5-13 | 1.30 |  |
|  |  |  | Female | 5-17 | 5.15 |  |
|  |  |  | Female | 5-9 | 0.25 |  |
|  |  | 2014 | Combined | 10-13 | 3.30 |  |
|  |  |  | Combined | 10-17 | 7.02 |  |
|  |  |  | Combined | 14-17 | 10.93 |  |
|  |  |  | Combined | 5-13 | 1.68 |  |
|  |  |  | Combined | 5-17 | 4.35 |  |
|  |  |  | Combined | 5-9 | 0.48 |  |
|  |  |  | Male | 10-13 | 3.26 |  |
|  |  |  | Male | 10-17 | 4.88 |  |
|  |  |  | Male | 14-17 | 6.57 |  |
|  |  |  | Male | 5-13 | 1.74 |  |
|  |  |  | Male | 5-17 | 3.15 |  |
|  |  |  | Male | 5-9 | 0.62 |  |
|  |  |  | Female | 10-13 | 3.35 |  |
|  |  |  | Female | 10-17 | 9.29 |  |
|  |  |  | Female | 14-17 | 15.59 |  |
|  |  |  | Female | 5-13 | 1.62 |  |
|  |  |  | Female | 5-17 | 5.63 |  |
|  |  |  | Female | 5-9 | 0.33 |  |
|  |  | 2015 | Combined | 10-13 | 3.65 |  |
|  |  |  | Combined | 10-17 | 7.85 |  |
|  |  |  | Combined | 14-17 | 12.35 |  |
|  |  |  | Combined | 5-13 | 1.87 |  |
|  |  |  | Combined | 5-17 | 4.86 |  |
|  |  |  | Combined | 5-9 | 0.54 |  |
|  |  |  | Male | 10-13 | 3.36 |  |
|  |  |  | Male | 10-17 | 5.31 |  |
|  |  |  | Male | 14-17 | 7.37 |  |
|  |  |  | Male | 5-13 | 1.86 |  |
|  |  |  | Male | 5-17 | 3.45 |  |
|  |  |  | Male | 5-9 | 0.75 |  |
|  |  |  | Female | 10-13 | 3.95 |  |
|  |  |  | Female | 10-17 | 10.55 |  |
|  |  |  | Female | 14-17 | 17.69 |  |
|  |  |  | Female | 5-13 | 1.87 |  |
|  |  |  | Female | 5-17 | 6.36 |  |
|  |  |  | Female | 5-9 | 0.32 |  |
|  |  | 2016 | Combined | 10-13 | 4.17 |  |
|  |  |  | Combined | 10-17 | 8.38 |  |
|  |  |  | Combined | 14-17 | 12.89 |  |
|  |  |  | Combined | 5-13 | 2.11 |  |
|  |  |  | Combined | 5-17 | 5.20 |  |
|  |  |  | Combined | 5-9 | 0.55 |  |
|  |  |  | Male | 10-13 | 3.77 |  |
|  |  |  | Male | 10-17 | 5.36 |  |
|  |  |  | Male | 14-17 | 7.04 |  |
|  |  |  | Male | 5-13 | 2.06 |  |
|  |  |  | Male | 5-17 | 3.50 |  |
|  |  |  | Male | 5-9 | 0.76 |  |
|  |  |  | Female | 10-13 | 4.59 |  |
|  |  |  | Female | 10-17 | 11.60 |  |
|  |  |  | Female | 14-17 | 19.20 |  |
|  |  |  | Female | 5-13 | 2.17 |  |
|  |  |  | Female | 5-17 | 7.02 |  |
|  |  |  | Female | 5-9 | 0.33 |  |
|  |  | 2017 | Combined | 10-13 | 4.33 |  |
|  |  |  | Combined | 10-17 | 8.69 |  |
|  |  |  | Combined | 14-17 | 13.33 |  |
|  |  |  | Combined | 5-13 | 2.23 |  |
|  |  |  | Combined | 5-17 | 5.45 |  |
|  |  |  | Combined | 5-9 | 0.63 |  |
|  |  |  | Male | 10-13 | 3.73 |  |
|  |  |  | Male | 10-17 | 5.58 |  |
|  |  |  | Male | 14-17 | 7.53 |  |
|  |  |  | Male | 5-13 | 2.09 |  |
|  |  |  | Male | 5-17 | 3.67 |  |
|  |  |  | Male | 5-9 | 0.83 |  |
|  |  |  | Female | 10-13 | 4.96 |  |
|  |  |  | Female | 10-17 | 12.03 |  |
|  |  |  | Female | 14-17 | 19.61 |  |
|  |  |  | Female | 5-13 | 2.39 |  |
|  |  |  | Female | 5-17 | 7.34 |  |
|  |  |  | Female | 5-9 | 0.42 |  |
|  |  | 2018 | Combined | 10-13 | 4.74 |  |
|  |  |  | Combined | 10-17 | 9.25 |  |
|  |  |  | Combined | 14-17 | 14.05 |  |
|  |  |  | Combined | 5-13 | 2.43 |  |
|  |  |  | Combined | 5-17 | 5.81 |  |
|  |  |  | Combined | 5-9 | 0.63 |  |
|  |  |  | Male | 10-13 | 4.33 |  |
|  |  |  | Male | 10-17 | 6.44 |  |
|  |  |  | Male | 14-17 | 8.67 |  |
|  |  |  | Male | 5-13 | 2.35 |  |
|  |  |  | Male | 5-17 | 4.20 |  |
|  |  |  | Male | 5-9 | 0.81 |  |
|  |  |  | Female | 10-13 | 5.17 |  |
|  |  |  | Female | 10-17 | 12.25 |  |
|  |  |  | Female | 14-17 | 19.83 |  |
|  |  |  | Female | 5-13 | 2.51 |  |
|  |  |  | Female | 5-17 | 7.53 |  |
|  |  |  | Female | 5-9 | 0.45 |  |
| Loikas 2013^a^ [] | Sweden | 2010 | Male | All ages | 15.35 | No dispensation in the previous 1 year |
|  |  |  | Female | All ages | 24.71 |  |
| ^a^Incidence per 1000 person-years.  ^b^Incidence per 1000 population. | | | | | | |
