## Supplementary material for "Age-sex specific prevalence and incidence of antidepressant prescribing in the Nordic countries: a systematic review": Table 1

| **Table 1.** Study characteristics of included studies. | | | | | | | | |
| --- | --- | --- | --- | --- | --- | --- | --- | --- |
| **Study** | **Setting/context** | **Country** | **Study period** | **Sample size** | **Ages** | **Female, %** | **Types of antidepressants** | **Outcomes reported** |
| Abbing-Karahagopian 2014 | Nationwide prescription registry | Denmark | 2001-2009 | Not specified | All | Not specified | All antidepressants | Prevalence |
| Autti-Rämö 2011 | Nationwide prescription registry | Finland | 1997-2007 | 1.7 million | 0-26 | 48.9 | All antidepressants | Prevalence and incidence |
| Bachmann 2016 | Nationwide prescription registry | Denmark | 2005-2012 | 1.2 million | 0-19 | 48.7 | All antidepressants | Prevalence |
| Bojanić 2024 | Nationwide prescription registries | Denmark, Norway, Sweden | 2006-2021 | Not specified | All | Not specified | All antidepressants | Prevalence |
| Corneliusson 2024 | Population-based clinical study | Sweden | 2000-2017 | 1611 | 85 and older | 60.9 to 82.8 | All antidepressants | Prevalence |
| Forslund 2020 | Regional administrative health care data | Sweden | 2007-2017 | 1.7 million | 20 and older | Not specified | All antidepressants excluding amitriptylin | Prevalence |
| Foulon 2010 | Nationwide prescription registry | Finland | 1998-2005 | Not specified | 0-19 | Not specified | All antidepressants | Prevalence and incidence |
| Gómez-Lumbreras 2021 | Nationwide prescription registries | Denmark, Norway, Sweden | 2008-2017 | Not specified | 0-19 | Not specified | All antidepressants | Prevalence |
| Hansen 2007 | Regional prescription database | Denmark | 1992-2004 | 0.5 million | 20 and older | Not specified | All antidepressants | Prevalence |
| Hartz 2016a | Nationwide prescription registry | Norway | 2004-2013 | Not specified | 13-17 | Not specified | All antidepressants | Prevalence and incidence |
| Hartz 2016b | Nationwide prescription registry | Norway | 2004-2014 | 1.1 million | 0-17 | 48.8 | All antidepressants | Prevalence |
| Ingemann 2021 | Population-based and nationwide medical registers | Denmark, including Greenland | 2019 | 5.2 million | 10-89 | 50.3 | All antidepressants | Prevalence |
| Ishtiak-Ahmed 2023 | Nationwide prescription registry | Denmark | 2015-2019 | 1.2 million | 65 and older | About 54 | All antidepressants | Incidence |
| Kjosavik 2011 | Nationwide prescription registry | Norway | 2004-2009 | 4.8 million | All | Not specified | All antidepressants | Incidence |
| Kjosavik 2009 | Nationwide prescription registry | Norway | 2005 | 4.6 million | All | Not specified | All antidepressants | Prevalence |
| Lagerberg 2019 | Nationwide prescription registry | Sweden | 2006-2013 | Not specified | 0-24 | Not specified | All antidepressants | Prevalence |
| Lien 2023 | Population-based survey linked to prescription registry | Norway | 2004-2020 | 0.1 million | 15-19 | About 50 | All antidepressants | Prevalence |
| Loikas 2013 | Nationwide prescription registry | Sweden | 2010 | 9.3 million | All | Not specified | All antidepressants | Prevalence and incidence |
| Pottegård 2014 | Nationwide prescription registry | Denmark | 1995-2011 | 0.8 million | 5-17 | Not specified | SSRI | Prevalence and incidence |
| Rasmussen 2024 | Nationwide prescription registries | Denmark, Norway, Sweden | 2007-2018 | 2.8 million | 5-17 | Not specified | All antidepressants | Incidence |
| Saastamoinen 2012 | Nationwide prescription registry | Finland | 1999-2005 | Not specified | 0-17 | Not specified | SSRI | Incidence |
| Sihvo 2008 | Population-based survey linked to nationwide prescription registry | Finland | 1999-2003 | 7112 | 30 and older | 52 | All antidepressants | Prevalence |
| Sihvo 2010 | Nationwide prescription registry | Finland | 1994-2003 | 4.1 million | 18 and older | Not specified | All antidepressants | Prevalence |
| Skovlund 2017 | Nationwide prescription registry | Denmark | 2000-2013 | 40.2 million person-years | 10-49 | Not specified | All antidepressants, excluding bupropion | Prevalence and incidence |
| Steffenak 2012 | Nationwide prescription registry | Norway | 2006-2010 | Not specified | 15-16 | Not specified | All antidepressants | Prevalence |
| Steinhausen 2014 | Nationwide prescription registry | Denmark | 1996-2010 | Not specified | 0-17 | Not specified | All antidepressants | Prevalence |
| Wastesson 2012 | Nationwide prescription registry | Sweden | 2008 | 0.5 million | 80 and older | 63.7 | All antidepressants | Prevalence |
| Wesselhoeft 2020 | Nationwide prescription registries | Denmark, Norway, Sweden | 2007-2017 | 3.7 million | 5-17 | Not specified | All antidepressants | Prevalence |
| Zito 2006 | Population-based prescription database | Denmark | 2000 | 0.5 million | 0-19 | Not specified | All antidepressants | Prevalence |
| Zoëga 2009 | Nationwide prescription registry | Iceland | 2003-2007 | Not specified | 0-17 | Not specified | All antidepressants | Prevalence and incidence |
